# Missense variants in *ANKRD11* cause KBG syndrome by impairment of stability or transcriptional activity of the encoded protein

**DOI:** 10.1101/2021.12.20.21267971

**Authors:** Elke de Boer, Charlotte W. Ockeloen, Rosalie A. Kampen, Juliet E. Hampstead, Alexander J.M. Dingemans, Dmitrijs Rots, Lukas Lütje, Tazeen Ashraf, Rachel Baker, Mouna Barat-Houari, Brad Angle, Nicolas Chatron, The DDD study, Anne-Sophie Denommé-Pichon, Orrin Devinsky, Christèle Dubourg, Frances Elmslie, Houda Zghal Elloumi, Laurence Faivre, Sarah Fitzgerald-Butt, David Geneviève, Jacqueline A.C. Goos, Benjamin M. Helm, Jiddeke M. van de Kamp, Usha Kini, Amaia Lasa-Aranzasti, Gaetan Lesca, Sally A. Lynch, Irene M.J. Mathijssen, Ruth McGowan, Kristin G. Monaghan, Sylvie Odent, Rolph Pfundt, Audrey Putoux, Jeroen van Reeuwijk, Gijs W.E. Santen, Erina Sasaki, Jesitha Sivanathan, Arthur Sorlin, Peter J. van der Spek, Alexander P.A. Stegmann, Sigrid M.A. Swagemakers, Irene Valenzuela, Eléonore Viora-Dupont, Antonio Vitobello, Stephanie M. Ware, Mathys Wéber, Christian Gilissen, Karen J. Low, Simon E. Fisher, Lisenka E.L.M. Vissers, Maggie M.K. Wong, Tjitske Kleefstra

## Abstract

**Purpose:** Although haploinsufficiency of *ANKRD11* is among the most common genetic causes of neurodevelopmental disorders, the role of rare *ANKRD11* missense variation remains unclear. We characterized the clinical, molecular and functional spectra of *ANKRD11* missense variants.

**Methods:** We collected clinical information of individuals with *ANKRD11* missense variants and evaluated phenotypic fit to KBG syndrome. We assessed pathogenicity of variants by *in silico* analyses and cell-based experiments.

**Results:** We identified 29 individuals with (mostly *de novo*) *ANKRD11* missense variants, who presented with syndromic neurodevelopmental disorders and were phenotypically similar to individuals with KBG syndrome caused by *ANKRD11* protein truncating variants or 16q24.3 microdeletions. Missense variants significantly clustered in Repression Domain 2. Cellularly, most variants caused reduced ANKRD11 stability. One variant resulted in decreased proteasome degradation and loss of ANKRD11 transcriptional activity.

**Conclusion:** Our study indicates that pathogenic heterozygous missense variants in *ANKRD11* cause the clinically recognizable KBG syndrome. Disrupted transrepression capacity and reduced protein stability each independently lead to ANKRD11 loss-of-function, consistent with haploinsufficiency. This highlights the diagnostic relevance of *ANKRD11* missense variants, but also poses diagnostic challenges, as the KBG-associated phenotype may be mild and inherited pathogenic *ANKRD11* (missense) variants are increasingly observed, warranting stringent variant classification and careful phenotyping.

## INTRODUCTION

KBG syndrome (MIM#148050) is an autosomal dominant neurodevelopmental disorder (NDD) typically characterized by mild intellectual disability (ID) or developmental delay (DD), macrodontia of upper central permanent incisors, mild skeletal anomalies, behavioral disturbances and distinctive craniofacial features [1-6]. Although KBG syndrome is considered a clinically recognizable syndrome with macrodontia as its most defining trait [6], there is considerable clinical variability and none of the KBG features is pathognomonic. Hence, despite being described as a clinical entity since 1975 [1], KBG syndrome was underdiagnosed before the introduction of exome sequencing [7]. The exact prevalence of KBG syndrome is not established, but it is thought to be a relatively common cause of genetic NDD, with the associated gene (*ANKRD11*) in the top 3 of mutated genes in NDD cohorts, accounting for 0.5-1% of diagnoses [8, 9].

KBG syndrome is caused by heterozygous protein truncating variants (PTVs) in *ANKRD11* (ankyrin repeat domain-containing protein 11; ANKRD11) or by 16q24.3 microdeletions encompassing (part of) *ANKRD11*. PTVs and microdeletions explain all cases in four previously described KBG cohorts [2-5], whereas in the general population, *ANKRD11* shows a strong constraint against loss-of-function variation (pLI 1; o/e 0.05 [0.02-0.11]; v2.1.1) [10]. Therefore, haploinsufficiency of *ANKRD11* is commonly accepted as the mechanism of pathogenicity for KBG syndrome [11]. This is supported by observations of reduced amounts of *ANKRD11* mRNA and protein when the gene contains a PTV [12], suggesting that variants trigger the nonsense-mediated decay (NMD) pathway [12], although PTVs leading to a (partial) escape from NMD have also been described [7, 13]. Also consistent with haploinsufficiency is the finding that ANKRD11 mutated with p.(Lys1347del) or p.(Leu2143Val) shows reduced transcriptional activity on the *p21* promotor in cell-based systems, and that this effect can be rescued by wildtype but not mutated ANKRD11 [14].

ANKRD11 is ubiquitously expressed and localizes mainly to the nucleus in a homogenous pattern. ANKRD11 is a crucial regulator of neuronal development [7, 15] that interacts with coactivators and corepressors of transcription [16], showing (co)regulatory effects on various sets of genes. These include genes encoding signaling molecules, chromatin remodelers and transcriptional regulators [15], controlling histone acetylation and gene expression during neural development. ANKRD11 contains three transcriptional regulatory domains: one activation domain and two repression domains. The repression domains, located at the N-terminus (RD1) and C-terminus of ANKRD11 (RD2), functionally outweigh the activation domain, as full-length ANKRD11 functions as a repressor of ligand-dependent transcription [17]. Interaction of ANKRD11 with other proteins and homodimerization are mediated through ankyrin repeats, located at the N-terminus [13]. The C-terminal part of ANKRD11, containing (predicted) destruction box motifs (D-boxes), was suggested to be critical for its degradation [13].

Whereas PTVs in *ANKRD11* are a well-recognized cause of KBG syndrome, the role of rare missense variants remains ambiguous. Contrary to what is seen for PTVs, constraint metrics based on the general population indicate that missense variants tend to be well-tolerated (Z-score -0.55; o/e 1.04 [1-1.08]) [10]. There are numerous entries of *ANKRD11* missense variants in ClinVar (access date 20-08-2021) but only ∼6% are classified as (likely) pathogenic, and almost half as variants of uncertain significance [18]. In the literature, ∼2.6% of (*de novo*) variants in *ANKRD11* are missense variants [19], listed in Table S1. Missense variants are reported with varying levels of evidence on pathogenicity, and functional studies have only been performed for p.(Leu2143Val), showing a loss-of-function effect [14]. The Yoda mutant mouse (C3H.Cg-Ankrd11Yod/H, p.(Glu2502Lys)), carries an *Ankrd11* missense variant and shows phenotypic overlap with core features of KBG syndrome, including reduced body size as well as craniofacial abnormalities such as shortened snouts with deformed nasal bones, wider skulls and failure of cranial sutures to close [20]. Additionally, Yoda mice show behavioral abnormalities reflective of cognitive dysfunction [15]. On the cellular level, the heterozygous Yoda variant causes similar cellular perturbations of abnormal neuronal precursor proliferation and localization of neurons as seen for Ankrd11 knockdown [15], suggestive of a loss-of-function mechanism. However, a dominant-negative mechanism has also been hypothesized to contribute to the Yoda mouse phenotype [13]. Ankrd11 was shown to mislocalize to the nucleolus, possibly resulting from diminished degradation [13]. As the N-terminal ankyrin repeats are unaffected by the variant, dimerization of wildtype and mutant Ankrd11 was hypothesized to result in decreased degradation of both proteins, potentially implicating such dominant-negative mechanism [13]. So far, a dominant-negative mechanism has not been confirmed in additional studies. In general, consequences of *ANKRD11* missense variants on clinical phenotypes and protein function are largely unknown.

We characterized genotypes, phenotypes and functional consequences associated with *ANKRD11* missense variants by describing a cohort of 29 individuals. Most individuals exhibit both the characteristic facial appearance as well as other KBG-associated features, fitting well within the clinical spectrum described for KBG syndrome. We show that missense variants in *ANKRD11* significantly cluster in the C-terminal RD2, with an overrepresentation of mutated arginine residues. Missense variants result in a loss of normal ANKRD11 function, either caused by reduced protein stability with normal or increased proteasome degradation, or caused by a loss of transrepression capacity with decreased proteasome degradation. Our studies did not confirm ANKRD11 mislocalization or a dominant-negative mechanism. Instead, we found that missense variants result in loss-of-function.

## MATERIALS AND METHODS

### Clinical and *in silico* characterization

Identification and clinical characterization of individuals, and *in silico* analyses of (likely) pathogenic *ANKRD11* missense variants are described in the Supplementary information. Variants were annotated to NM_013275.6 in GRCh37/Hg19. Human Phenotype Ontology (HPO)-based [21] clustering was performed as previously described [22] using clinical data of 29 individuals with *ANKRD11* missense variants (Table S2, Supplementary JSON), and 30 individuals with *ANKRD11* PTVs or microdeletions, after grouping HPO-data based on semantic similarity (Table S3) [23, 24]. Spatial clustering of independently observed missense variants (25/29) was performed as previously described [25], excluding four familial variants. The Supplementary information contains brief details of both clustering analyses. *p*-values <0.05 were considered significant. Figure 2, Table S2, Figure S1 and Supplementary JSON are not included in this preprint and are available from the corresponding author on request. Consent procedures and details of the IRB/oversight body that provided approval or exemption for the research described are described in the ethical statement.

### Missense permutation analysis

To test whether the observed number of variants affecting arginine residues was significantly greater than expected by chance, *ANKRD11* (ENST00000301030.10_4/NM_013275.6) was mutated *in silico* and output was annotated with Ensembl Variant Effect Predictor (VEP) v104 [26]. To generate an expected missense distribution, sets of 17 missense variants in RD2 and 8 missense variants outside RD2 (based on 25 independently observed missense variants, excluding four familial variants) were randomly sampled 100,000 times using per-nucleotide mutation rates as weights [27]. The number of missense variants affecting arginine residues inside and outside RD2 were counted per iteration. *p*-values were computed using a permutation test, by ranking the observed number of variants affecting arginine residues within the set of 100,000 expected values. *p*-values <0.05 were considered significant.

### Cell culture and transfection

HEK293T/17 cells (CRL-11268, ATCC) were cultured in DMEM (Gibco) supplemented with 10% foetal bovine serum (Gibco) and 100□U/ml Penicillin-Streptomycin (Thermo Fisher) at 37°C and 5% CO2. For immunofluorescence analysis, cells were seeded onto coverslips coated with 100 μg/ml poly-D-lysine (Merck, Millipore). Transfections were performed using GeneJuice (Merck, Millipore) following the manufacturer’s instructions or polyethyenimine (PEI) in a 3:1 ratio with the total mass of DNA transfected.

### DNA Constructs and site-directed mutagenesis

Full-length wildtype ANKRD11 construct fused to a C-terminal Myc-DDK tag under a human CMV promoter (pCMV-Entry-ANKRD11) was purchased from Origene (RC211717). To generate an N-terminal EGFP-tag, sequence encoding EGFP was cut from a pEGFP-C2 vector (Clontech) and subcloned into the pCMV-Entry-ANKRD11 plasmid using KpnI/NdeI restriction sites. Constructs (pCMV-Entry-EGFP-ANKRD11) carrying (in frame) *ANKRD11* missense variants were generated using a QuikChange Lightning (Multi)Site-Directed Mutagenesis Kit (Agilent) following the manufacturer’s protocol. All constructs were verified by Sanger sequencing. Table S4 lists all primer sequences.

### Fluorescence imaging of subcellular localization

HEK 293T/17 cells grown on poly-D-lysine-coated coverslips were transiently transfected with 500ng of pCMV-EGFP-ANKRD11 constructs 24 hours after seeding. Cells were fixated 48 hours after transfection using 4% paraformaldehyde solution (Electron Microscopy Supplies Ltd) for 20 minutes at room temperature. Hoechst 33342 (Invitrogen) was used for nuclear staining, before mounting with VECTASHIELD® Antifade Mounting Medium (Vectorlab). Fluorescence images were obtained using an LSM880 AxioObserved confocal microscope (Zeiss). For images of single nuclei, the Airyscan unit (Zeiss) was used with a 4.0 zoom factor. Images were analyzed with the ImageJ “Analyze particle” plugin.

### Fluorescence-based quantification of protein stability and degradation

HEK293/T17 cells were transfected in triplicate in clear-bottomed black 96-well plates with EGFP-tagged ANKRD11 variants. After 48 hours, cycloheximide (Sigma) at 50μg/ml or MG132 (R&D Systems) at 5µg/ml was added. Cells were incubated at 37°C with 5% CO2 in the Infinite M200PRO microplate reader (Tecan), and fluorescence intensity of EGFP (Ex: 503nm, Em: 540nm) was measured over 24 hours at 3-hour intervals. Statistical analysis was done using two-way analysis of variance (ANOVA) followed by Dunnett’s correction for multiple testing. *p*-values <0.05 were considered significant.

### Luciferase reporter assays

We used firefly luciferase reporters: pGL2-p21 promoter-Luc and WWP-Luc carrying the promoter region of *CDKN1A*/*P21*. Reporters were gifts from Martin Walsh (Addgene plasmid #33021; http://n2t.net/addgene:33021; RRID:Addgene_33021) [28] and Bert Vogelstein (Addgene plasmid #16451; http://n2t.net/addgene:16451; RRID:Addgene_16451) [29] respectively. HEK293/T17 cells were seeded in clear-bottomed white 96-well plates (Greiner Bio-One). Cells were co-transfected with 320ng of firefly luciferase reporter construct, 6.5ng of pGL4.74 Renilla luciferase normalization control, and 1000ng of an EGFP-ANKRD11 expression construct or empty EGFP expression vector. After 48 hours, firefly luciferase and Renilla luciferase activities were measured using a Dual-Luciferase Reporter Assay system (Promega) according to the manufacturer’s instructions at an Infinite F Plex Microplate reader (Tecan). Statistical analysis was done using a one-way ANOVA followed by Dunnett’s *post-hoc* test. *p*-values <0.05 were considered significant.

## RESULTS

### *ANKRD11* missense variants cause syndromic neurodevelopmental phenotypes

Through international collaborations [30, 31] we identified 29 individuals with rare missense variants in *ANKRD11* (Figure 1; Table S2). The cohort consisted of 24 unrelated individuals with *ANKRD11* missense variants – of which 20 variants occurred *de novo* – and one family with five affected individuals from three generations (Figure S1). For five individuals, inheritance status of the variant could not be established. The cohort comprises 18 males and 11 females, with an age range of seven months to 73 years. We observed syndromic neurodevelopmental phenotypes, summarized in Table 1 with detailed data compiled in Table S2 and the Supplementary Information. Most frequent phenotypic features were facial dysmorphisms (28/29, 96.6%; 23/29 fitting characteristic dysmorphisms of KBG, 79.3%; Figure 2A), behavioral disturbances (25/28, 89.3%), neurodevelopmental delay (26/28, 92.9%; speech delay 23/26, 88.5%; motor delay 20/27, 74.1%), mild to moderate ID (22/27, 81.5%; borderline 2/27, 7.4%; mild 15/27, 55.6%; moderate 4/27, 14.8%, unknown severity 3/27, 11.1%), problems of dentition (21/26, 80.8%; macrodontia of upper central incisors 13/24, 54.2%; other dental abnormalities 14/21, 66.7%; Figure 2B) and hand abnormalities (20/25, 80%; Figure 2C). Of note, individual 19 also carried a likely pathogenic variant in *ARID2*, implicated in Coffin-Siris syndrome 6 (MIM#617808), and in the family of five individuals an additional *ANKRD11* variant of uncertain significance, p.(Pro61Ser), was observed in cis with the pathogenic p.(Arg2579His) variant.

**Table 1:**
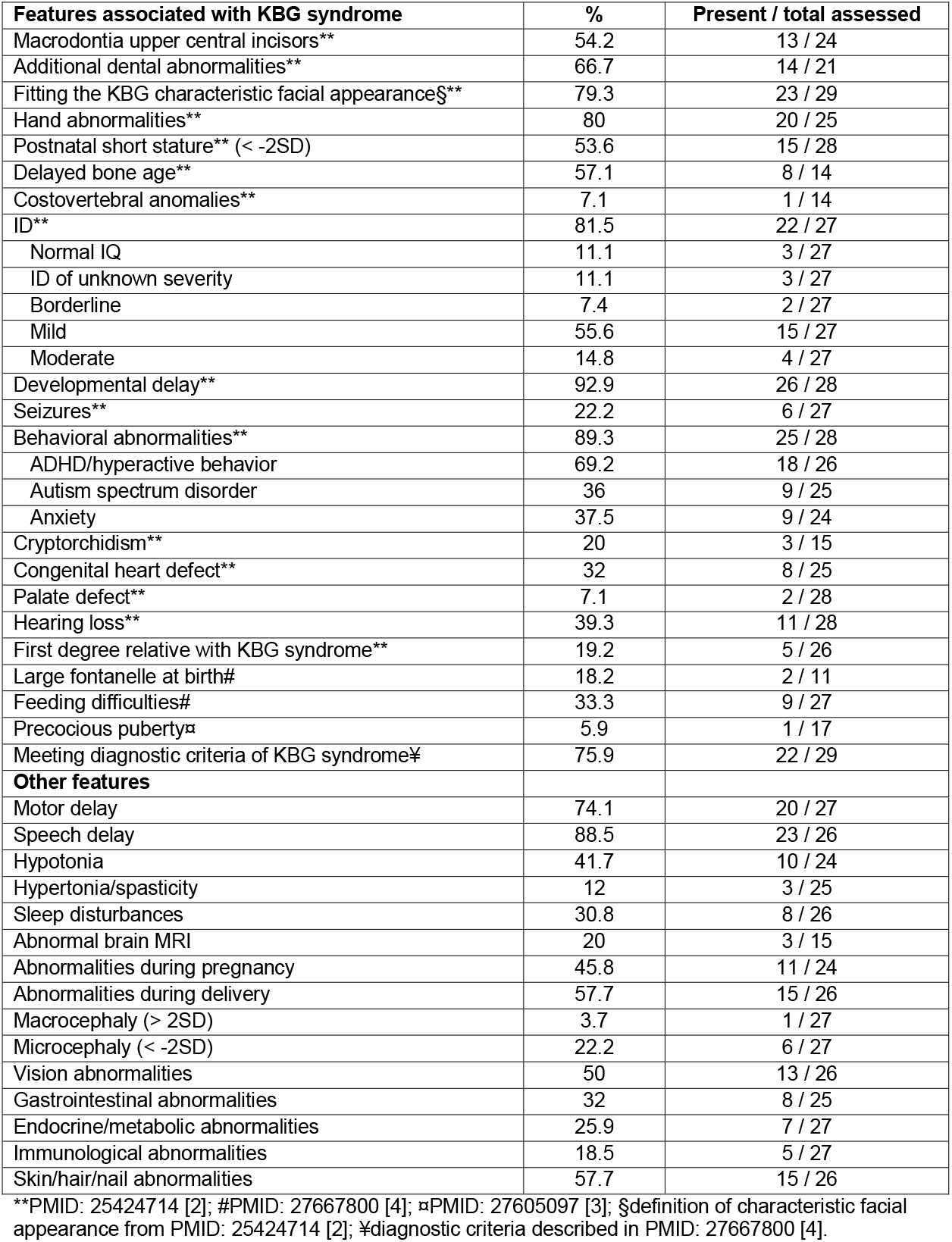
Summary of observed clinical features.

| **Features associated with KBG syndrome** | **%** | **Present / total assessed** |
| --- | --- | --- |
| Macrodontia upper central incisors** | 54.2 | 13 / 24 |
| Additional dental abnormalities** | 66.7 | 14 / 21 |
| Fitting the KBG characteristic facial appearance§** | 79.3 | 23 / 29 |
| Hand abnormalities** | 80 | 20 / 25 |
| Postnatal short stature** (< -2SD) | 53.6 | 15 / 28 |
| Delayed bone age** | 57.1 | 8 / 14 |
| Costovertebral anomalies** | 7.1 | 1 / 14 |
| ID** | 81.5 | 22 / 27 |
| Normal IQ | 11.1 | 3 / 27 |
| ID of unknown severity | 11.1 | 3 / 27 |
| Borderline | 7.4 | 2 / 27 |
| Mild | 55.6 | 15 / 27 |
| Moderate | 14.8 | 4 / 27 |
| Developmental delay** | 92.9 | 26 / 28 |
| Seizures** | 22.2 | 6 / 27 |
| Behavioral abnormalities** | 89.3 | 25 / 28 |
| ADHD/hyperactive behavior | 69.2 | 18 / 26 |
| Autism spectrum disorder | 36 | 9 / 25 |
| Anxiety | 37.5 | 9 / 24 |
| Cryptorchidism** | 20 | 3 / 15 |
| Congenital heart defect** | 32 | 8 / 25 |
| Palate defect** | 7.1 | 2 / 28 |
| Hearing loss** | 39.3 | 11 / 28 |
| First degree relative with KBG syndrome** | 19.2 | 5 / 26 |
| Large fontanelle at birth# | 18.2 | 2 / 11 |
| Feeding difficulties# | 33.3 | 9 / 27 |
| Precocious puberty▫ | 5.9 | 1 / 17 |
| Meeting diagnostic criteria of KBG syndrome¥ | 75.9 | 22 / 29 |
| **Other features** |  |  |
| Motor delay | 74.1 | 20 / 27 |
| Speech delay | 88.5 | 23 / 26 |
| Hypotonia | 41.7 | 10 / 24 |
| Hypertonia/spasticity | 12 | 3 / 25 |
| Sleep disturbances | 30.8 | 8 / 26 |
| Abnormal brain MRI | 20 | 3 / 15 |
| Abnormalities during pregnancy | 45.8 | 11 / 24 |
| Abnormalities during delivery | 57.7 | 15 / 26 |
| Macrocephaly (> 2SD) | 3.7 | 1 / 27 |
| Microcephaly (< -2SD) | 22.2 | 6 / 27 |
| Vision abnormalities | 50 | 13 / 26 |
| Gastrointestinal abnormalities | 32 | 8 / 25 |
| Endocrine/metabolic abnormalities | 25.9 | 7 / 27 |
| Immunological abnormalities | 18.5 | 5 / 27 |
| Skin/hair/nail abnormalities | 57.7 | 15 / 26 |

\*\*PMID: 25424714 [2]; #PMID: 27667800 [4]; ▫PMID: 27605097 [3]; §definition of characteristic facial appearance from PMID: 25424714 [2]; ¥diagnostic criteria described in PMID: 27667800 [4].

**Figure 1:**
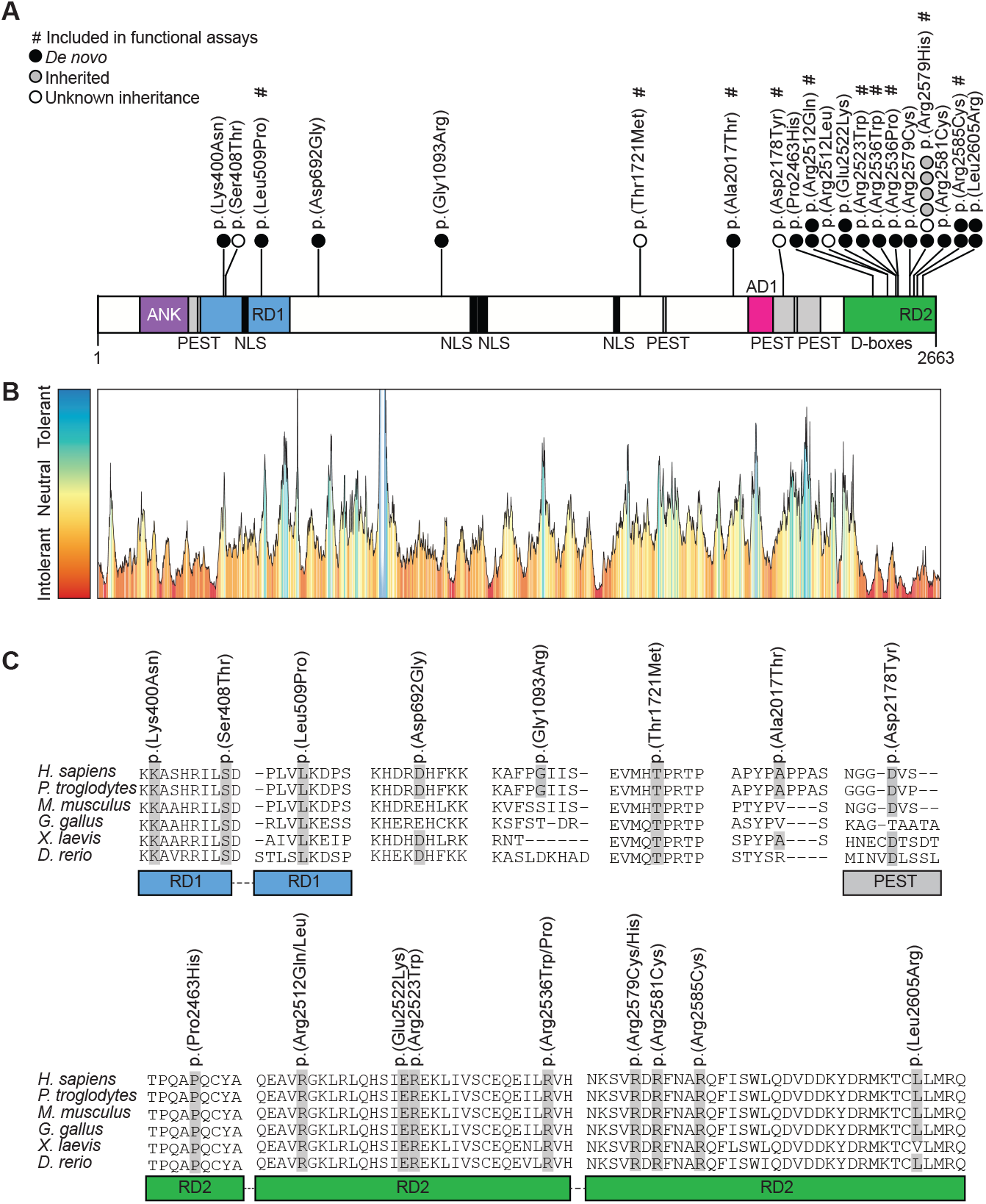
Missense variants cluster in the intrinsic repressor domain 2 in the C-terminus of the ANKRD11 protein. (A) Schematic representation of ANKRD11 (UniProt: Q6UB99) indicating the location of variants included in this study. *De novo* variants are indicated by black discs, inherited variants by grey discs, variants with unknown inheritance by white discs, and variants marked with # are included in functional assays. The ANKRD11 protein sequence (2663 amino acids) contains an ankyrin-repeat domain (ANK; purple; amino acids 133-296), four PEST sequences (grey; amino acids 286-324, 1796-1806, 2146-2212 and 2222-2297), four bipartite nuclear localization signals (NLS; black; amino acids 1184-1200, 1208-1236, 1358-1374, 1640-1656), two intrinsic repressor domains (RD1; blue; amino acids 318-611 and RD2; green; amino acids 2369-2663), and an activator domain (AD1; pink; amino acids 2076–2145). An overview with variant details per subject is provided in Table S2, with details on variant interpretation in Table S5. (B) MetaDome analysis of the ANKRD11 missense variants. Overview of the ANKRD11 protein (NM_013275.5/ NP_037407.4) tolerance landscape visualized via the MetaDome web server version 1.0.1. The green and blue peaks correspond to regions more tolerant to missense variation, and the red valleys indicate intolerant regions. (C) Sequence alignment of the region containing part of ANKRD11 amino acid sequence in human (UniProt: Q6UB99), chimpanzee (A0A2I3TR65), mouse (E9Q4F7), chicken (A0A3Q2UE98), African clawed frog (A0A1L8GEN1) and zebrafish (E7F5R3). Residues in which missense variants were found are highlighted in grey.

**Figure 2:** Clinical evaluation of individuals with *ANKRD11* missense variants. Figure 2 is not included in this preprint. Images are available from the corresponding author on request.

### *ANKRD11* missense variants are predicted to be deleterious and cluster at C-terminal RD2

We found 20 unique missense variants (Figure 1; Table S5), significantly clustering at the highly intolerant C-terminal RD2 (*p*=9.99e-9), with recurrence of p.(Arg2512Gln), p.(Glu2522Lys), p.(Arg2579His), p.(Arg2585Cys) and p.(Leu2605Arg). Additionally, the arginine residues at p.2512, p.2536 and p.2579 were affected by two different missense variants, and p.(Glu2522Lys) is equivalent to the orthologous *Ankrd11* p.(Glu2502Lys) in the Yoda mouse (Figure S2). Most of the observed variants are predicted to be damaging, and affect conserved and intolerant residues (Figure 1B-C), with no predicted effects on pre-mRNA splicing (Table S5). Two variants, p.(Glu2522Lys) and p.(Arg2523Trp), affect a ProViz [32] predicted (low consensus similarity) D-box (Table S6, Figure S3), and two variants, p.(Arg2512Leu) and p.(Arg2512Gln), are located at an additional RxxL-motif, marked as D-box in prior literature [13]. We observed a significantly greater number of missense variants affecting arginine residues within RD2 (12/17 variants; 70.6%) than would be expected by chance (*p*=1.00e-4), while such enrichment was not observed outside RD2 (0/8 variants; 0%; *p*=0.37), visualized in Figure S4. We therefore hypothesized that generally, *ANKRD11* missense variants affecting arginine residues in RD2 are likely to be pathogenic.

### Phenotypes associated with *ANKRD11* missense variants fit the KBG-associated clinical spectrum

Based on four large published cohorts together describing 135 individuals with KBG syndrome caused by PTVs or 16q24.3 microdeletions affecting *ANKRD11* [2-5], we assessed the phenotypic fit of each of the individuals in the missense cohort to the clinical spectrum associated with KBG syndrome. The majority of individuals (23/29, 79.3%) exhibited dysmorphisms fitting the characteristic facial gestalt by which KBG syndrome can be recognized (Figure 2A-B; Table S2, Table S5). Indeed, two individuals in the cohort (individual 6 and 14) were diagnostically evaluated by targeted *ANKRD11* Sanger sequencing because of a high clinical suspicion. Of the 29 individuals, 22 (75.9%) met the diagnostic criteria described for KBG syndrome [4] (Table S2). After review of the observed phenotypes by expert clinicians, it was concluded that almost all individuals fit the KBG-associated phenotypic spectrum. These included many individuals not fully meeting the diagnostic criteria, either because some features of KBG syndrome not captured in the diagnostic criteria (e.g. delayed bone age and congenital heart defects) were seen in these individuals, or because the characteristic facial appearance of KBG syndrome was observed. Only for individuals 3 and 29, carrying p.(Leu509Pro) and p.(Leu2605Arg), phenotypic fit to the clinical spectrum of KBG syndrome was considered poor.

We next compared the group of individuals with missense variants to the collective KBG cohorts [2-5] and found that frequencies of most KBG-associated features observed in the missense cohort lie within the range of frequencies of these features seen in the group of individuals with PTVs or microdeletions (Table 2). We therefore hypothesized that KBG syndrome resulting from missense variants is indistinguishable from KBG syndrome caused by *ANKRD11* PTVs or microdeletions.

**Table 2:** Comparison of clinical features between individuals with *ANKRD11* missense variants and previously reported KBG cohorts.

| Publication | Ockeloen et al.** |  | Low et al.# |  | Goldenberg et al.▣ |  | Gnazzo et al.¶ |  | Cumulative frequencies in published cohorts**,#,▣,¶ |  |  | This publication |  |
| --- | --- | --- | --- | --- | --- | --- | --- | --- | --- | --- | --- | --- | --- |
| Variant type | PTVs + microdeletions |  | PTVs |  | PTVs + microdeletions |  | PTVs + microdeletions |  | PTVs + microdeletions |  |  | Missense variants |  |
| Features associated with KBG syndrome | % | Present / total assessed | % | Present / total assessed | % | Present / total assessed | % | Present / total assessed | % | Present / total assessed | Frequency range (%) | % | Present / total assessed |
| Macrodonia upper central incisors** | 75.8 | 25 / 33 | 85.2 | 23 / 27 | 69.2 | 18 / 26 | 76.7 | 23 / 30 | 76.7 | 89 / 116 | 69.2-85.2 | 54.2 | 13 / 24 |
| Additional dental abnormalities** | 48.5 | 16 / 33 | NA | NA | NA | NA | NA | NA | 48.5 | 16 / 33 | 48.5 | 66.7 | 14 / 21 |
| Fitting characteristic facial appearance of KBG§** | 93.9 | 31 / 33 | 40.6 | 13 / 32 | 100 | 39 / 39 | 100 | 31 / 31 | 84.4 | 114 / 135 | 40.6-100 | 79.3 | 23 / 29 |
| Hand abnormalities** | 81.8 | 27 / 33 | 46.9 | 15 / 32 | 69.7 | 23 / 33 | 67.7 | 21 / 31 | 66.7 | 86 / 129 | 46.9-81.1 | 80 | 20 / 25 |
| Postnatal short stature** | 54.5 | 18 / 33 | 28.1 | 9 / 32 | 40.5 | 15 / 37 | 58.1 | 18 / 31 | 45.1 | 60 / 133 | 28.1-58.1 | 53.6 | 15 / 28 |
| Delayed bone age** | 15.2 | 5 / 33 | 60 | 3 / 5 | 66.7 | 8 / 12 | NA | NA | 32 | 16 / 50 | 15.2-66.7 | 57.1 | 8 / 14 |
| Costovertebral anomalies** | 30.3 | 10 / 33 | NA | NA | NA | NA | NA | NA | 30.3 | 10 / 33 | 30.3 | 7.1 | 1 / 14 |
| ID** | 60.6 | 20 / 33 | NA | NA | 60 | 9 / 15 | 59.1 | 13 / 22 | 60 | 42 / 70 | 59.1-60.6 | 81.5 | 22 / 27 |
| Learning disability or developmental delay** | 93.9 | 31 / 33 | 100 | 32 / 32 | 94.3 | 33 / 35 | 96.8 | 30 / 31 | 96.2 | 126 / 131 | 93.9-100 | 92.9 | 26 / 28 |
| Seizures** | 27.3 | 9 / 33 | 43.8 | 14 / 32 | 31.6 | 12 / 38 | 16.1 | 5 / 31 | 29.9 | 40 / 134 | 16.1-43.8 | 22.2 | 6 / 27 |
| Behavioral abnormalities** | 78.8 | 26 / 33 | 100 | 30 / 30 | 51.4 | 19 / 37 | NA | NA | 75 | 75 / 100 | 51.4-100 | 89.3 | 25 / 28 |
| Cryptorchidism** | 40 | 8 / 20 | 31.3 | 5 / 16 | 18.8 | 3 / 16 | 13.3 | 2 / 15 | 26.9 | 18 / 67 | 13.3-40 | 20 | 3 / 15 |
| Congenital heart defect** | 18.2 | 6 / 33 | 12.5 | 4 / 32 | 25.6 | 10 / 39 | 35.5 | 11 / 31 | 23 | 31 / 135 | 12.5-35.5 | 32 | 8 / 25 |
| Palate defect** | 21.2 | 7 / 33 | 12.5 | 4 / 32 | 5.1 | 2 / 39 | 0 | 0 / 31 | 9.6 | 13 / 135 | 0-21.2 | 7.1 | 2 / 28 |
| Hearing loss** | 24.2 | 8 / 33 | 25 | 8 / 32 | 30.6 | 11 / 36 | 19.4 | 6 / 31 | 25 | 33 / 132 | 19.4-30.6 | 39.3 | 11 / 28 |
| First degree relative with KBG syndrome** | 52.6 | 10 / 19 | 21.9 | 7 / 32 | 20.5 | 8 / 39 | 6.5 | 2 / 31 | 22.3 | 27 / 121 | 6.5-52.6 | 19.2 | 5 / 26 |
| Large fontanelle at birth# | NA | NA | 21.9 | 7 / 32 | NA | NA | NA | NA | 21.9 | 7 / 32 | 21.9 | 18.2 | 2 / 11 |
| Feeding difficulties# | NA | NA | 31.3 | 10 / 32 | NA | NA | 35.5 | 11 / 31 | 33.3 | 21 / 63 | 31.3-35.3 | 33.3 | 9 / 27 |
| Precocious puberty▣ | NA | NA | NA | NA | 15.6 | 5 / 32 | 5.6 | 1 / 18 | 12 | 6 / 50 | 5.6-15.6 | 5.9 | 1 / 17 |

\*\*PMID: 25424714 [2]; #PMID: 27667800 [4]; ▣PMID: 27605097 [3]; ¶PMID: 32124548 [5]; §definition of characteristic facial appearance from PMID: 25424714 [2].

To quantitatively investigate this hypothesis, we compared standardized clinical data (Supplementary JSON [21]) of 29 individuals with missense variants and 30 individuals with KBG syndrome caused by PTVs or 16q24.3 microdeletions affecting *ANKRD11*. Applying a Partitioning Around Medoids clustering algorithm [33] on 68 features derived from HPO-data resulted in correct classification of 37 of 59 individuals as either belonging to the PTV or missense variant group (p=0.04536; Figure 2D; Table S3A-B), challenging the clinical observation that pathogenic *ANKRD11* missense variants and PTVs or microdeletions lead to the same clinical entity.

### *ANKRD11* missense variants act via two distinct loss-of-function mechanisms

We continued by studying the functional consequences on protein localization, protein stability, proteasome degradation and transcriptional activity of a subset of the observed *ANKRD11* missense variants using HEK293T/17 cells transiently transfected with mutant ANKRD11. To obtain a comprehensive insight in the spectrum of variants, we examined variants spread across *ANKRD11*, including p.(Leu509Pro) located in RD1, p.(Thr1721Met), p.(Ala2017Thr) and p.(Asp2178Tyr) outside known protein domains, and several variants in RD2, being p.(Arg2512Gln), p.(Arg2523Trp), p.(Arg2536Trp), p.(Arg2536Pro), p.(Arg2579His) and p.(Arg2585Cys).

We first assessed the impact of missense variants on subcellular localization of ANKRD11. When transiently expressed as EGFP-fusion proteins in HEK239T/17 cells, wildtype ANKRD11 localized to the nucleus in a homogeneous speckle-like pattern, consistent with previous findings in non-neuronal cell lines [16] and mouse neocortical neurons overexpressing wildtype Ankrd11 [7]. None of the tested missense variants affected the nuclear localization of ANKRD11 (Figure 3, Figure S5), contrasting with the nucleolar mislocalization of mutant Ankrd11 that has been reported for Yoda mice [13].

**Figure 3:**
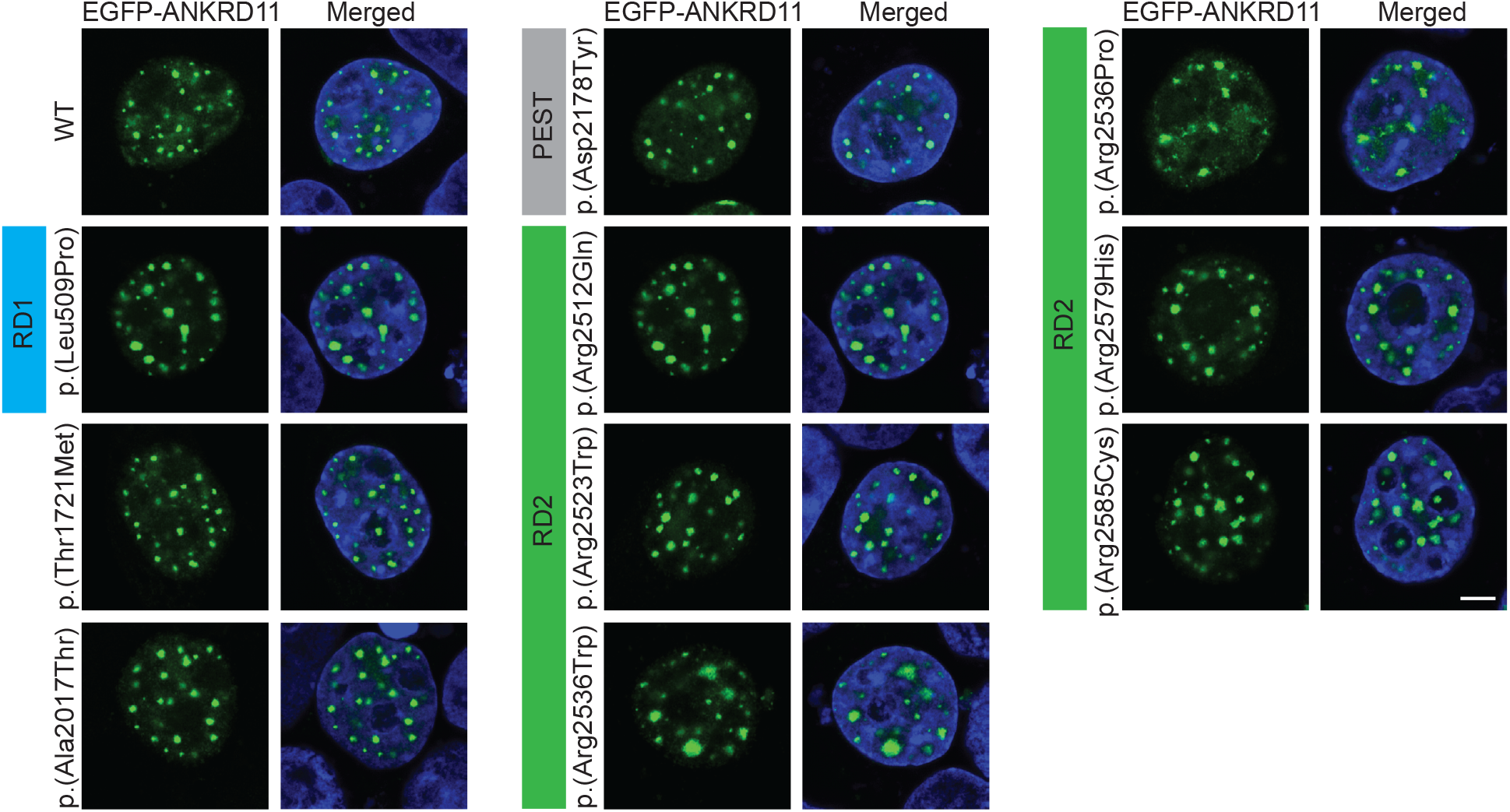
ANKRD11 variants do not affect its subcellular localization in transiently transfected HEK293T/17 cells. (A) Direct fluorescence imaging of cells expressing EGFP-tagged variants of the ANKRD11 protein using confocal microscopy. Wildtype and all variants showed a speckle-like pattern in the nucleus. Nuclei are stained with Hoechst 33342 (blue). Protein domains in which variants are located are indicated. Results are representative of three independent experiments. Scale bar= 5μm.

Based on spatial clustering at the C-terminus, which is critical for ANKRD11 degradation [13], we hypothesized that missense variants might alter ANKRD11 stability, possibly via altered proteasome degradation. Moreover, four variants are located at putative destruction motifs (Table S6, Figure S3). To assess ANKRD11 protein stability, we treated HEK239T/17 cells expressing EGFP-tagged ANKRD11 with cycloheximide (CHX) to inhibit translation, and measured relative fluorescence intensity over 24 hours. We found that all variants in RD2 except p.(Arg2585Cys) demonstrated reduced protein stability compared to wildtype, whereas among variants outside the RD2, only p.(Ala2017Thr) was less stable (Figure 4A, Figure S6-S7). All other variants outside RD2 showed similar stability to wildtype. We next examined the effect of missense variants on proteasome-mediated degradation after treating EGFP-ANKRD11 expressing cells with proteasome inhibitor MG132. Only two variants, both located in RD2, affected proteasome degradation, showing opposite directions of effect: while p.(Arg2523Trp) displayed increased proteasome degradation, p.(Arg2585Cys) demonstrated decreased proteasome degradation (Figure 4B).

**Figure 4:**
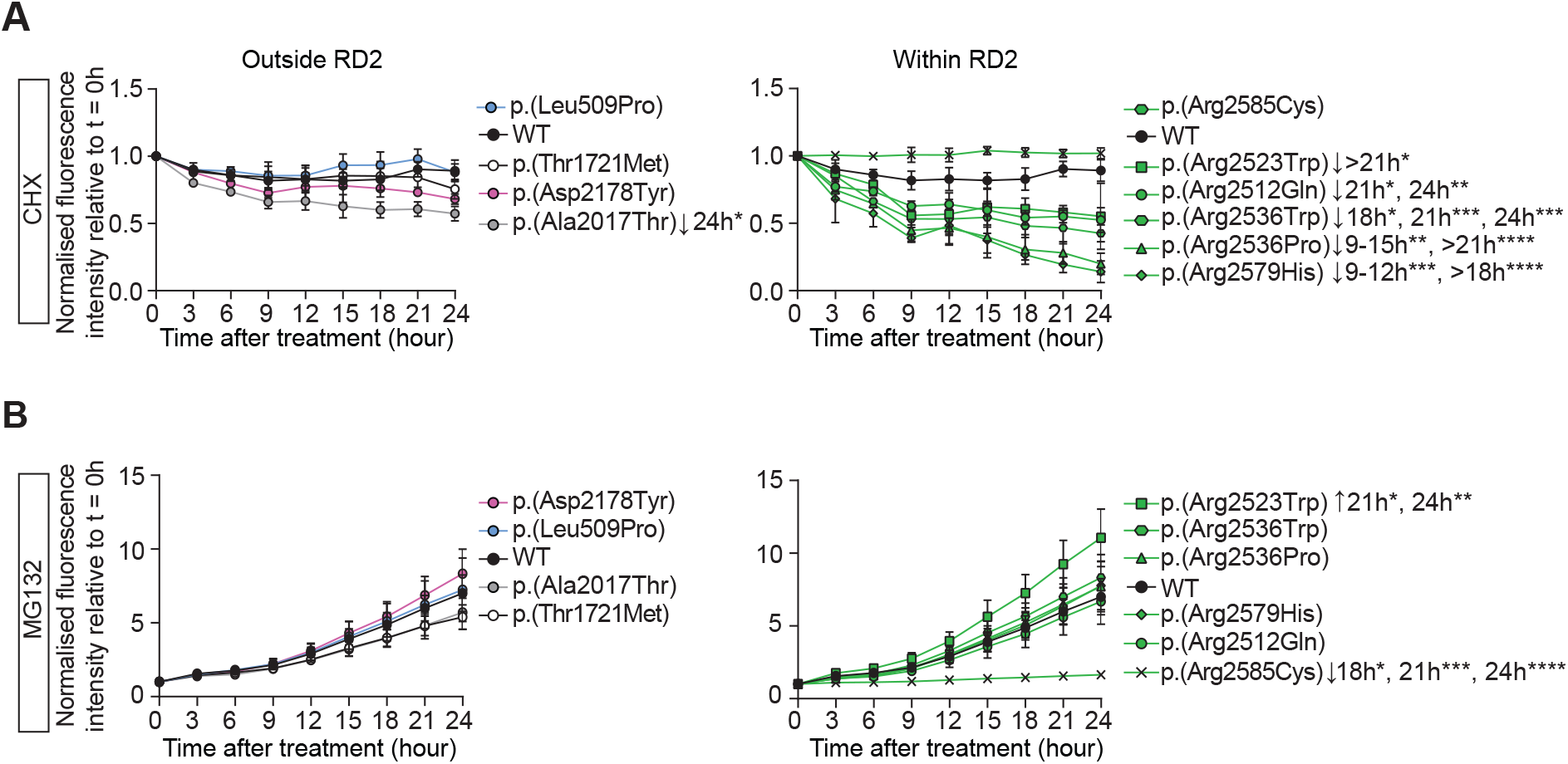
Reduced protein stability and impaired proteasome degradation of ANKRD11 variants in RD2 domain. (A) Relative fluorescence intensity of EGFP-tagged ANKRD11 variants overexpressed in HEK293T/17 cells treated with translation inhibitor cycloheximide (CHX; 50µg/ml). Equal volume of DMSO was used as a vehicle control. Fluorescence intensity was measured for 24 hours with three-hour intervals. Values are expressed relative to t= 0 hour and represent the mean ± SEM of three independent experiments, each preformed in triplicates (*p< 0.05, **p< 0.01, ***p< 0.001, **** p< 0.0001; two-way ANOVA and a post-hoc Dunnett’s test). (B) Relative fluorescence intensity of EGFP-tagged ANKRD11 variants overexpressed in HEK293T/17 cells treated with proteasome inhibitor MG132 (5µg/ml). Equal volume of DMSO was used as a vehicle control. Fluorescence intensity was measured for 24 hours with three-hour intervals. Values are expressed relative to t= 0 hour and represent the mean ± SEM of three independent experiments, each preformed in triplicates (*p< 0.05, **p< 0.01, ***p< 0.001, **** p< 0.0001; two-way ANOVA and a post-hoc Dunnett’s test).

To study the effects of missense variation on transcriptional activity of ANKRD11, we performed luciferase reporter assays with the *CDKN1A/P21* promoter, a known downstream target. Of all tested variants, only p.(Arg2585Cys) affected transcriptional activity, leading to a loss of transcriptional repression on *CDKN1A/P21* (Figure 5).

**Figure 5:**
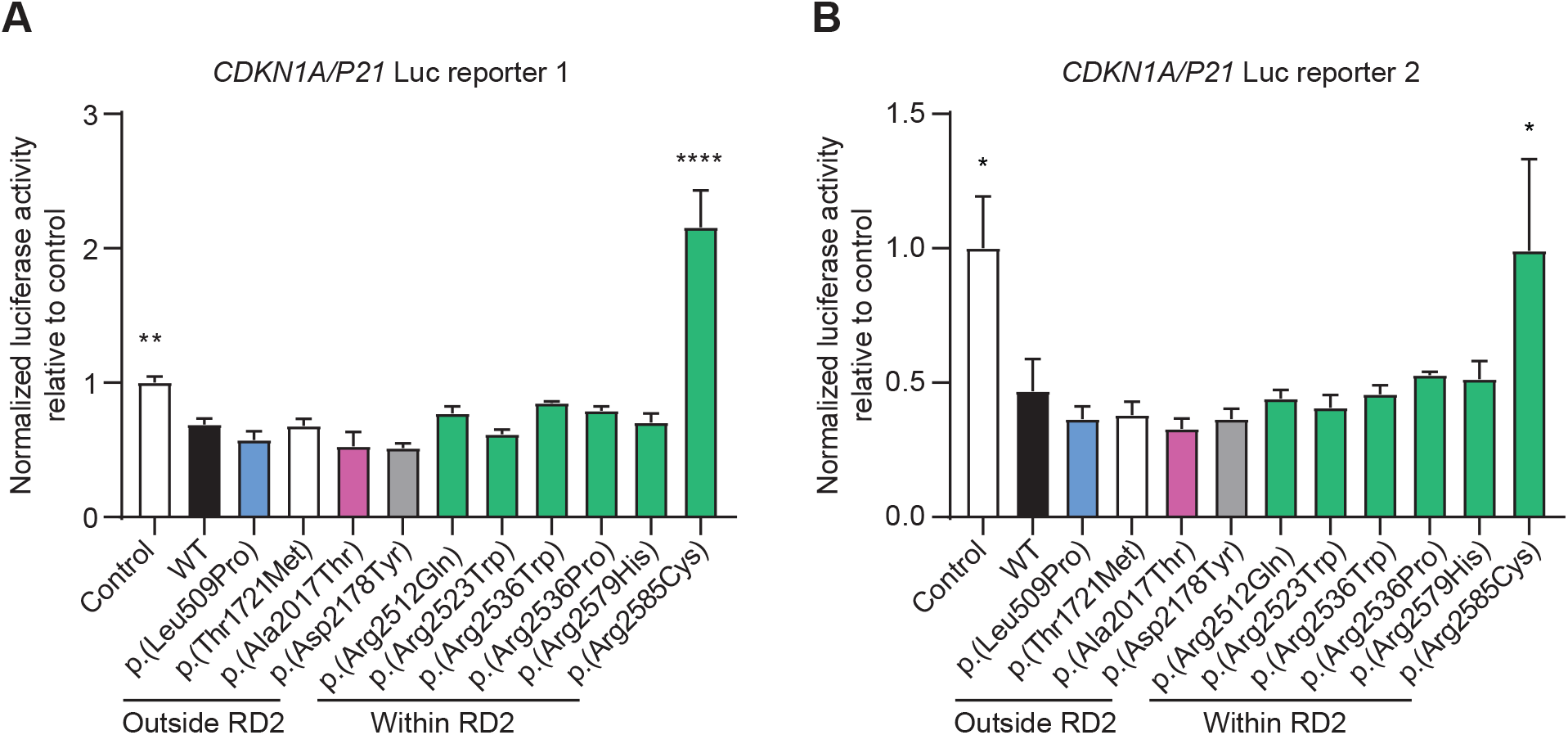
Loss of CDKN1A/P21 transcription repression caused by an ANKRD11 variant in RD2 domain. Results of luciferase assay with constructs containing WT and ANKRD11 variants, and two firefly luciferase reporter constructs with a CDKN1A/P21 promoter. Values are expressed relative to the control condition that used a EGFP-C2 construct without ANKRD11 and represent the mean ± SEM of three independent experiments, each performed in triplicate (*p<0.05, **p<0.01, ****p<0.0001 versus WT; one-way ANOVA and a post-hoc Dunnett’s test).

Taken together, our cell-based assays indicate that most missense variants yield a loss-of-function effect, either resulting from a reduced dosage of ANKRD11 due to decreased protein stability with or without increased proteasome degradation, or through a loss of transrepressive activity. Three variants, p.(Leu509Pro), p.(Thr1721Met) and p.(Asp2178Tyr), did not result in aberrations in any of the tested protein functions, and are therefore classified as of uncertain significance, although the individuals carrying p.(Thr1721Met) and p.(Asp2178Tyr) exhibit typical KBG features (individual 6 and 8, respectively). A summary of all evidence per observed missense variant including classification based on ACMG criteria is provided in Table 3 and Table S5.

**Table 3:** Classification of pathogenicity for the observed *ANKRD11* missense variants.

| g.DNA (Hg19/GRCh37) | cDNA (NM_013275.6) | Protein Effect | ACMG classification | Individual(s) |
| --- | --- | --- | --- | --- |
| Chr16:89371659G>A | c.181C>T | p.(Pro61Ser) | Uncertain Significance | 20, 21, 22, 23, 24 |
| Chr16:89351750C>G | c.1200G>C | p.(Lys400Asn) | Likely Pathogenic | 1 |
| Chr16:89351728A>T | c.1222T>A | p.(Ser408Thr) | Uncertain Significance | 2 |
| Chr16:89351424A>G | c.1526T>C | p.(Leu509Pro) | Uncertain Significance | 3 |
| Chr16:89350875T>C | c.2075A>G | p.(Asp692Gly) | Likely Pathogenic | 4 |
| Chr16:89349673C>T | c.3277G>A | p.(Gly1093Arg) | Likely Pathogenic | 5 |
| Chr16:89347788G>A | c.5162C>T | p.(Thr1721Met) | Uncertain Significance | 6 |
| Chr16:89346901C>T | c.6049G>A | p.(Ala2017Thr) | Pathogenic | 7 |
| Chr16:89346418C>A | c.6532G>T | p.(Asp2178Tyr) | Uncertain Significance | 8 |
| Chr16:89345562G>T | c.7388C>A | p.(Pro2463His) | Likely Pathogenic | 9 |
| Chr16:89341535C>T | c.7535G>A | p.(Arg2512Gln) | Pathogenic | 10, 11 |
| Chr16:89341535C>A | c.7535G>T | p.(Arg2512Leu) | Likely Pathogenic | 12 |
| Chr16:89341506C>T | c.7564G>A | p.(Glu2522Lys) | Likely Pathogenic | 13, 14 |
| Chr16:89341503G>A | c.7567C>T | p.(Arg2523Trp) | Pathogenic | 15 |
| Chr16:89341329G>A | c.7606C>T | p.(Arg2536Trp) | Pathogenic | 16 |
| Chr16:89341328C>G | c.7607G>C | p.(Arg2536Pro) | Pathogenic | 17 |
| Chr16:89337296G>A | c.7735C>T | p.(Arg2579Cys) | Pathogenic | 18 |
| Chr16:89337295C>T | c.7736G>A | p.(Arg2579His) | Pathogenic | 19, 20, 21, 22, 23, 24 |
| Chr16:89337290G>A | c.7741C>T | p.(Arg2581Cys) | Likely Pathogenic | 25 |
| Chr16:89337278G>A | c.7753C>T | p.(Arg2585Cys) | Pathogenic | 26, 27 |
| Chr16:89335064A>C | c.7814T>G | p.(Leu2605Arg) | Pathogenic | 28, 29 |

## DISCUSSION

Although KBG syndrome has been clinically recognized for almost 50 years [1], and PTVs and microdeletions affecting *ANKRD11* have been robustly implicated in its etiology since 2011 [7], the role of rare missense variation in *ANKRD11* remained unclear. We characterized the clinical, molecular and functional spectra of *ANKRD11* missense variants, by collecting information for 29 individuals and assessing effects of missense variation on ANKRD11 functions. We show that almost all individuals carrying rare heterozygous predicted damaging *ANKRD11* missense variants fit well within the clinical spectrum described for KBG syndrome. Missense variants mainly affect the C-terminal RD2 with an overrepresentation of mutated arginine residues. Based on cellular assays, missense variants result in loss-of-function of ANKRD11, either by impaired protein stability or reduced transcriptional activity, consistent with *ANKRD11* haploinsufficiency causing KBG syndrome through PTVs and microdeletions.

Most individuals presented with characteristics fitting the KBG-associated phenotypic spectrum, and from a clinical perspective, individuals with KBG syndrome caused by *ANKRD11* missense variants or by PTVs or microdeletions are indistinguishable. However, unexpectedly, HPO-based clustering analysis showed a statistically significant difference between the groups. Possibly, ascertainment bias influenced this analysis, as recognizing pathogenicity for missense variants is more challenging than for PTVs. Additionally, out of seven individuals with missense variants that did not meet the KBG diagnostic criteria, six were correctly assigned to the missense cluster (Table S3B), potentially (in part) driving the observed difference. Lastly, the cohort of cases with PTVs was obtained from one expert health care centre, whereas the missense cohort represents an international collaboration. Larger cohorts are needed to assess whether there are indeed phenotypic differences, or whether these results can be explained by cohort effects.

The clinical variability of KBG syndrome is noteworthy, showing considerable phenotypic differences between affected individuals within the same family or between unrelated individuals with the same variant. This variability is best illustrated by comparing individuals 13 and 14, carrying *de novo* p.(Glu2522Lys). Although both presented with macrodontia and the characteristic facial appearance, individual 13 exhibited moderate ID, behavioral disturbances, hypotonia, a duplex kidney, strabismus and normal growth, whereas individual 14 had normal intelligence, no neurobehavioral abnormalities, a submucous cleft palate, moderate hearing loss, mild growth hormone deficiency and microcephaly. Also the family with five affected individuals is a key example, in whom two presented with macrodontia (individual 21 and 24) and three exhibited the characteristic facial gestalt (individual 20, 23 and 24). We therefore argue not to rule out pathogenicity for individual *ANKRD11* missense variants on inheritance or clinical grounds only. Also for the two individuals (individual 3 and 29) not clearly exhibiting symptoms of KBG syndrome, the variants are classified as variant of uncertain significance and pathogenic variant when applying ACMG criteria alone (Table S5; p.(Leu509Pro) and p.(Leu2605Arg) respectively) [34].

*ANKRD11* shows significant regional differences in missense depletion in the general population, with three distinct regions of missense tolerance: p.1-p.415 with modest regional missense depletion (o/e 0.51), p.416-p.2276 tolerating missense variation (o/e 1.1) and p.2277-p.2664 showing high missense depletion (o/e 0.11) [35]. Consistently, in our cohort we observed that variants significantly clustered in the highly depleted C-terminal region, which was previously suggested to be implicated in the mechanism underlying the KBG phenotype [13], although we also observed missense variants in the tolerant middle and N-terminal depleted regions. The proportion of independently mutated arginine residues is remarkable (total cohort 12/25, 48%; RD2 12/17, 70.6%), and more pronounced than the overrepresentation of mutated arginine residues seen for pathogenic variants underlying genetic disorders in general (15-20% [36, 37]). Arginine is also the most frequently mutated residue in all secondary structures when considering pathogenic variants [37]. Therefore, we hypothesize that the molecular underpinnings of the observed overrepresentation of mutated arginine residues lies in the three-dimensional structure of ANKRD11, which could not be taken into account, as the crystal structure of ANKRD11 is largely uncharacterized and *ab initio* models are unreliable, despite recent advances in the field [38]. However, based on our *in silico* studies, we argue that if missense variants in *ANKRD11* affect an arginine residue in the C-terminal RD2, this is suggestive for pathogenicity.

Regarding functional impact, the majority of tested missense variants resulted in reduced protein stability, but it was only for p.(Arg2523Trp) that this could be explained by increased proteasome degradation. We hypothesize that variants reducing protein stability without impairment of proteasome degradation affect other mechanisms implicated in protein homeostasis that could be activated by ubiquitination (e.g. autophagy). Of note, p.(Arg2523Trp) is located at a putative D-box possibly affecting ANKRD11 ubiquitination and subsequent proteasome degradation. However, the other tested variant at a D-box, p.(Arg2512Gln), showed no impairment of proteasome degradation, which challenges the previous suggestion that disruption of the C-terminal D-boxes is the sole pathophysiological mechanism of KBG syndrome [13]. In contrast, the only other variant with altered proteasome degradation, p.(Arg2585Cys), showed reduced proteasome degradation, and might slightly increase protein stability, although the latter was not statistically significant. The p.(Arg2585Cys) variant is also the only tested variant that resulted in reduced transcriptional repression on *CDKN1A/P21*, consistent with previous observations for p.(Leu2143Val) [14]. These findings suggest that p.(Arg2585Cys) results in loss-of-function, despite a potential accumulation of mutant ANKRD11 that contrasts with the dosage reduction seen for the other tested variants. Lastly, we did not observe changes in ANKRD11 subcellular localization for the assessed missense variants, contrarily to what has been reported for Yoda mice with p.(Glu2502Lys) [13].

The three variants that did not show aberrations in any of our assays are all located outside RD2, and classified as variants of uncertain significance (Table S5). Of the four tested variants located outside RD2, only p.(Ala2017Thr) affected the assessed protein functions, whereas all tested variants located inside RD2 did affect protein function. It is therefore possible that variants outside RD2 exert effects on ANKRD11 functions that were not captured by our studies. Alternatively, they might alter pre-mRNA splicing (Figure S8) [39] despite low SpliceAI scores [40]. Based on the role of ANKRD11 in chromatin-remodelling, evaluating transcriptomic and epigenetic profiles of individuals or cell models could help to increase understanding of the effects of missense variants in the various domains.

In conclusion, our study shows that (*de novo*) pathogenic missense variants in *ANKRD11* cause the clinically recognizable KBG syndrome, with a similar phenotypic spectrum as previously observed for PTVs and microdeletions affecting *ANKRD11*. We demonstrate that loss of transrepression capacity and reduced protein stability are independent molecular mechanisms by which missense variants cause a functional loss of ANKRD11. These findings add to the mechanistic complexity underlying *ANKRD11* haploinsufficiency, already comprising deletion of the locus [3], putative null alleles [12], and PTVs escaping the NMD pathway [7, 13], although effects of the latter on protein stability and function have not yet been elucidated. Because inheritance of pathogenic variants in *ANKRD11* is regularly observed due to variability of the associated phenotype, missense variants present with diagnostic challenges, warranting stringent variant classification and careful phenotyping. However, as KBG syndrome is a relatively common cause of genetic NDD, the involvement of *ANKRD11* missense variants in cohorts of undiagnosed individuals with NDD should be considerable.

## Data availability

Code used for spatial clustering is shared at https://github.com/laurensvdwiel/SpatialClustering. Code used for permutation testing is available at https://github.com/jhampstead/ANKRD11-simulations. For missense cases, all available phenotypic information in HPO terminology is shared as a supplementary file (Supplementary JSON). The HPO-data of individuals obtained from the Radboudumc Biobank are not publicly available due to IRB and General Data Protection Regulation (EU GDPR) restrictions. Access to these data may be requested from the data availability committee by contacting the corresponding author. Model and code for HPO-based clustering analysis are available at https://github.com/ldingemans/HPO_clustering_Wang. Figure 2, Table S2, Figure S1 and Supplementary JSON are not included in this preprint and are available from the corresponding author on request.

## Data Availability

https://github.com/jhampstead/ANKRD11-simulations

https://github.com/ldingemans/HPO_clustering_Wang

## Acknowledgements

We are very grateful to all individuals and their families for their participation in this study. This work was financially supported by Aspasia grants of the Dutch Research Council (015.014.036 to TK and 015.014.066 to LV), Netherlands Organization for Health Research and Development (91718310 to TK), and the Max Planck Society (MW, SEF). Individual 4 was sequenced in the Scottish Genomes Partnership. The Scottish Genomes Partnership was funded by the Chief Scientist Office of the Scottish Government Health Directorates [SGP/1] and The Medical Research Council Whole Genome Sequencing for Health and Wealth Initiative (MC/PC/15080). In addition, the collaborations in this study were facilitated by ERN ITHACA, one of the 24 European Reference Networks (ERNs) approved by the ERN Board of Member States, co-funded by European Commission. The aims of this study contribute to the Solve-RD project (EdB, ASDP, LF, CG, TK, AV, LV) which has received funding from the European Union’s Horizon 2020 research and innovation programme under grant agreement No 779257.

## Author Information

CO, MW and TK designed the study. Clinical data collection and interpretation was performed by EdB, CO and TK. AD carried out HPO-based clustering analysis. JH and CG performed the mutational clustering and missense permutation analysis. MW designed cell-based experiments, which were executed by RK and MW. Cellular images were quantitatively investigated with code written by LL. EdB, CO, DR, TA, RB, MBH, BA, NC, ASDP, OD, CD, FE, HZE, LF, SFB, DG, JG, BH, JK, UK, ALA, GL, SL, IM, RMG, KZM, SO, RP, AP, JvR, GS, ES, JS, AS, PS, APAS, SS, IV, EVD, AV, SW, MWe, KL and TK participated in recruitment of individuals, phenotyping and/or next-generation sequencing analysis. EdB, MW, CO, LV, SEF and TK analyzed and interpreted the results. EdB, MW, CO and TK wrote the manuscript. CO, MW, and TK supervised the project. All authors contributed to the final version of the manuscript.

## Ethics Declaration

We obtained informed consent to publish unidentifiable data for all individuals reported in this study. Specific consent was obtained for publication of clinical photographs. Consent procedures were in accordance with the Declaration of Helsinki and local ethical guidelines of the participating centres. The institutional review board ‘Commissie Mensgebonden Onderzoek Regio Arnhem-Nijmegen’ approved this study under number 2011/188. This number refers to performing diagnostic exome sequencing. Discovery of novel syndromes and description of clinical cohorts from this series can be taken as such. All the appropriate institutional forms have been archived locally.

## Notes

**Disclosure** HZE and KZM are employees of GeneDx, Inc. There are no other conflicts of interest.

### Competing Interest Statement

HZE and KZM are employees of GeneDx, Inc.

### Funding Statement

This study was funded by Aspasia grants of the Dutch Research Council 015.014.036 to TK and 015.014.066 to LV, Netherlands Organization for Health Research and Development 91718310 to TK, and the Max Planck Society MW, SEF. Individual 4 was sequenced in the Scottish Genomes Partnership. The Scottish Genomes Partnership was funded by the Chief Scientist Office of the Scottish Government Health Directorates SGP 1 and The Medical Research Council Whole Genome Sequencing for Health and Wealth Initiative MC PC 15080. In addition, the collaborations in this study were facilitated by ERN ITHACA, one of the 24 European Reference Networks ERNs approved by the ERN Board of Member States, co-funded by European Commission. The aims of this study contribute to the Solve-RD project EdB, ASDP, LF, CG, TK, AV, LV which has received funding from the European Union Horizon 2020 research and innovation programme under grant agreement No 779257.

### Author Declarations

*Ethics committee/IRB Commissie Mensgebonden Onderzoek Regio Arnhem-Nijmegen* of *institution name Radboudumc* gave ethical approval for this work under number 2011/188

