## Supplementary Figures S1-S8 for "Missense variants in *ANKRD11* cause KBG syndrome by impairment of stability or transcriptional activity of the encoded protein"

**Figure S1: Pedigree of the family with five individuals carrying p.(Arg2579His)**

Not included in this preprint. Available from the corresponding author on request.

#### Figure S2: Variant p.(Glu2522Lys) in individuals 13 and 14 is equivalent to Yoda variant p.(Glu2502Lys)

Alignment of human ANKRD11 (Q6UB99) and mouse Ankrd11 (E9Q4F7) using CLUSTAL O(1.2.4) multiple sequence alignment. De human p.(Glu2522Lys) variant and Yoda mouse variant p.(Glu2502Lys) are marked.

```
SP|Q6UB99|ANR11_HUMAN MPKGGCPKAPQQEELPLSSDMVEKQTGKKDKDKVSLTKTPKLERGDGGKEVRERASKRKL 60
SP|E9Q4F7|ANR11_MOUSE MPKGGCSKTPQQEDFALSNDMVEKQTGKKDKDKVSLTKTPKLDRSDGGKEVRERATKRKL 60
                      ****** *:****:: **.***********************:*.**********:****

SP|Q6UB99|ANR11_HUMAN PFTAGANGEQKDSDTEKQGPERKRIKKEPVTRKAGLLFGMGLSGIRAGYPLSERQQVALL 120
SP|E9Q4F7|ANR11_MOUSE PFTVGANGEQKDSDTEKQGPERKRIKKEPVARKSGLLFGMGLSGIRAGYPLSERQQVALL 120
                      ***.**************************:**:**************************

SP|Q6UB99|ANR11_HUMAN MQMTAEESANSPVDTTPKHPSQSTVCQKGTPNSASKTKDKVNKRNERGETRLHRAAIRGD 180
SP|E9Q4F7|ANR11_MOUSE MQMTAEESANSPVDTTPKHPSQSTVCQKGTPNSASKTKDKVNKRNERGETRLHRAAIRGD 180
                      ************************************************************

SP|Q6UB99|ANR11_HUMAN ARRIKELISEGADVNVKDFAGWTALHEACNRGYYDVAKQLLAAGAEVNTKGLDDDTPLHD 240
SP|E9Q4F7|ANR11_MOUSE ARRIKELISEGADVNVKDFAGWTALHEACNRGYYDIAKQLLAAGAEVNTKGLDDDTPLHD 240
                      ***********************************:************************

SP|Q6UB99|ANR11_HUMAN AANNGHYKVVKLLLRYGGNPQQSNRKGETPLKVANSPTMVNLLLGKGTYTSSEESSTESS 300
SP|E9Q4F7|ANR11_MOUSE AANNGHYKVVKLLLRYGGNPQQSNRKGETPLKVANSPTMVNLLLGKGTYTSSEESSTESS 300
                      ************************************************************

SP|Q6UB99|ANR11_HUMAN EEEDAPSFAPSSSVDGNNTDSEFEKGLKHKAKNPEPQKATAPVKDEYEFDEDDEQDRVPP 360
SP|E9Q4F7|ANR11_MOUSE EEEDAPSFAPSSSVDGNNTDSEFEKGLKLKAKNPEPQKTVTPVKDEYEFDEDDEQDRVPP 360
                      **************************** *********:.:*******************

SP|Q6UB99|ANR11_HUMAN VDDKHLLKKDYRKETKSNSFISIPKMEVKSYTKNNTIAPKKASHRILSDTSDEEDASVTV 420
SP|E9Q4F7|ANR11_MOUSE VDDKHLLKKDYRKEAKANSFISIPKMEVKSYSKNNTLAPKKAAHRILSDTSDEEDVSVSI 420
                      **************:*:**************:****:*****:************.**::

SP|Q6UB99|ANR11_HUMAN GTGEKLRLSAHTILPGSKTREPSNAKQQKEKNKVKKKRKKETKGREVRFGKRSDKFCSSE 480
SP|E9Q4F7|ANR11_MOUSE GAGEKLRLSAHTMLPGSKARESSSSRQQKEKNKLKKKRKKETKGKEVRFGKRSDKFCSSG 480
                      *:**********:*****:** *.::*******:**********:**************

SP|Q6UB99|ANR11_HUMAN SESESSESGEDDRDSLGSSGCLKGSPLVLKDPSLFSSLSASSTSSHGSSAAQKQNPSHTD 540
SP|E9Q4F7|ANR11_MOUSE SESESSESEEDDGDSVGSSGCLKGSPLVLKDPSLFSSLSASSTSSHGSAVAQKHGSGHTD 540
                      ******** *** **:********************************:.***:. .***

SP|Q6UB99|ANR11_HUMAN QHTKHWRTDNWKTISSPAWSEVSSLSDSTRTRLTSESDYSSEGSSVESLKPVRKRQEHRK 600
SP|E9Q4F7|ANR11_MOUSE QHTKHWRTDNWKAISSPAWSEVSSLSDSSRTGLTSESDCSSEGSSVESLKPTRRKQEHRK 600
                      ************:***************:** ****** ************.*::*****

SP|Q6UB99|ANR11_HUMAN RASL----SEKKSPFLSSAEGAVPKLDKEGKVVKKHKTKHKHKNKEKGQCSISQELKLKS 656
SP|E9Q4F7|ANR11_MOUSE RGVLQSAPSEKRSSFHPCTDGAVPKLDKEGKVVKKHKTKHKHKHKEKGQCSVSQELKLKS 660
                      *. *    ***:* *  .::***********************:*******:********

SP|Q6UB99|ANR11_HUMAN FTYEYEDSKQKSDKAILLENDLSTENKLKVLKHDRDHFKKEEKLSKMKLEEKEWLFKDEK 716
SP|E9Q4F7|ANR11_MOUSE FTYEYEDSKQKSDKAILLESDLSTENKLKVLKHDREHLKKEDKLGRMKPEDKDWLFKDEK 720
                      *******************.***************:*:***:**.:** *:*:*******

SP|Q6UB99|ANR11_HUMAN SLKRIKDTNKDISRSFREEKDRSNKAEKERSLKEKSPKEEKLRLYKEERKKKSKDRPSKL 776
SP|E9Q4F7|ANR11_MOUSE VLKRIKDANKDMSRAFREDKDRASKAERERATKDKSPKEEKLRLYKEERKKKSKDRASRL 780
                       ******:***:**:***:***:.***:**: *:********************** *:*

SP|Q6UB99|ANR11_HUMAN EKKNDLKEDKISKEKEKIFKEDKEKLKKEKVYREDSAFDEYCNKNQFLENEDTKFSLSDD 836
SP|E9Q4F7|ANR11_MOUSE ERKNDMKEDKLSKEKEKAFKEDKEKLKKEKLYREDAAFDDYCNKSQFLDHEDTKFSLSDD 840
                      *:***:****:****** ************:****:***:****.***::**********

SP|Q6UB99|ANR11_HUMAN QRDRWFSDLSDSSFDFKGEDSWDSPVTDYRDMKSDSVAKLILETVKEDSKERRRDSRARE 896
SP|E9Q4F7|ANR11_MOUSE QQERWFSDLSDSSFDFKGEDSWDS-VTDYRDIKNDSVAKLILETVKEDSKEKKRDNKIRE 899
                      *::********************* ******:*.*****************::**.: **
```

```
SP|Q6UB99|ANR11_HUMAN KRDYREPFFRKKDRDYLDKNSEKRKEQTEKHKSVPGYLSEKDKKRRESAEAGRDRKDALE 956
SP|E9Q4F7|ANR11_MOUSE KRDFKDSFFRKRDRDCLDKNSEKRRDQTEKHKSIPSYLSEKDKKRRESAEGGRDR----- 954
                      ***::: ****:*** ********::*******:*.**************.****

SP|Q6UB99|ANR11_HUMAN SCKERRDGRAKPEEAHREELKECGCESGFKDKSDGDFGKGLEPWERHHPAREKEKKDGPD 1016
SP|E9Q4F7|ANR11_MOUSE -----RDGRIRSEEVHREDLKECGFESSFKDKSDCDFPKNLEPWERPHAAREKEKKDALE 1009
                           **** : **.***:***** **.****** ** *.****** * ********. :

SP|Q6UB99|ANR11_HUMAN KERKEKTKPERYKEKSSDKDKSEKSILEKCQKDKEFDKCFKEKKDTKEKHKDTHGKDKER 1076
SP|E9Q4F7|ANR11_MOUSE KERKEKGRADKYKEKSSERERSDKSTLDKCQKDKEFEKCFKEKKDGKEKHKDIHSKD--R 1067
                      ****** : ::******::::*:** *:********:******** ****** *.**  *

SP|Q6UB99|ANR11_HUMAN KASLDQGKEKKEKAFPGIISEDFSEKKDDKKGKEKSWYIADIFTDESEDDRDSCMGSGFK 1136
SP|E9Q4F7|ANR11_MOUSE KASFDQLREKKEKVFSSIISEDFSERKDDRKGKEKSWYIADIFTDESEDEKDDCVAGSFK 1127
                      ***:** :*****.* .********:***:******************::*.*:...**

SP|Q6UB99|ANR11_HUMAN MGEASDLPRTDGLQEKEEGREAYASDRHRKSSDKQHPERQKDKEPRDRRKDRGAADAGRD 1196
SP|E9Q4F7|ANR11_MOUSE ATEASDTQRVDGLPEKEEGREHPSDRHRKSSSDRQHTEKPRDKEPKEKKKDRGASEGGKD 1187
                        ****  *.*** *******  :. :::.***:** *: :****:::*****::.*:*

SP|Q6UB99|ANR11_HUMAN KK---EKVFEKHKEKKDKESTEKYKDRKDRASVDSTQDKKNKQKLPEKAEKKHAAEDKAK 1253
SP|E9Q4F7|ANR11_MOUSE KKEKMEKIFEKHKEKKDKECAERYKDRKERASADSAPEKKNKQKLPEKVEKKHFAEDKVK 1247
                      **   **:***********.:*:*****:***.**: :**********.**** ****.*

SP|Q6UB99|ANR11_HUMAN SKHKEKSDKEHSKE--RKSSRSADAEKSLLEKLEEEALHEYREDSNDKISEVSSDSFTDR 1311
SP|E9Q4F7|ANR11_MOUSE SKHKEKPEKEHSREREKPSRGPDVEKSLLEKLEEEALHDYREDSNDKISEVSSDSFADH 1307
                      ****** :****:*  ** **. *.**************:*****************:*:

SP|Q6UB99|ANR11_HUMAN GQEPGLTAFLEVSFTEPPGDDKPRESACLPEKLKEKERHRHSSSSSKKSHDRERAKKEKA 1371
SP|E9Q4F7|ANR11_MOUSE GQEPSLSTLLEVSFSEPPAEDKARDSACLSEKLREKERHRHSSSSSKKSHERERAKKEKA 1367
                      ****.*:::*****:***.:** *:**** ***:****************:*********

SP|Q6UB99|ANR11_HUMAN EKKEKGEDYKEG--GSRKDSGQYEKDFLEADAYGVSYNMKADIEDELDKTIELFSTEKKD 1429
SP|E9Q4F7|ANR11_MOUSE EKKEKSEDYKDSISSVRKDASQFEKDFLDAETYGVSYPTKADVEEELDKAIELFSSEKKD 1427
                      *****.****:.  . ***:.*:*****:*::*****  ***:*:****:*****:****

SP|Q6UB99|ANR11_HUMAN KNDSEREPSKKIEKELKPYGSSAINILKEKKKREKHREKWRDEKERHRDRHADGLLRHHR 1489
SP|E9Q4F7|ANR11_MOUSE RSDPEREPAKRIEKELKPYGSSAISILKEKKKREKHRERWREEKERHRDKHVDGFLRHH- 1486
                      :.* ****:*:*************.************:**:*******:*.**:****

SP|Q6UB99|ANR11_HUMAN DELLRHHRDEQKPATRDKDSPPRVLKDKSRDEGPRLGDAKLKEKFKDGAEKEKGDPVKMS 1549
SP|E9Q4F7|ANR11_MOUSE -------KDEPKPAAKDKDNPPNSFKEKSREESLKLSETKLKEKFKENTEREKGDSIKMS 1539
                             :** ***::***.**. :*:***:*. :*.::*******:.:*:**** :***

SP|Q6UB99|ANR11_HUMAN NGNDKVAPSKDPGKKDARPREKLLGDGDLMMTSFERMLSQKDLEIEERHKRHKERMKQME 1609
SP|E9Q4F7|ANR11_MOUSE NGNDKLVPSRDSGKKDSRPREKLLGDGDLMMTSFERMLSQKDLEIEERHKRHKERMKQME 1599
                      *****:.**:* ****:*******************************************

SP|Q6UB99|ANR11_HUMAN KLRHRSGDPKLKEKAKPADDGRKKGLDIPAKKPPGLDPPFKDKKLKESTPIPPAAENKLH 1669
SP|E9Q4F7|ANR11_MOUSE KMRHRSGDPKLKEK-KPTEDGRKKSLDFPSKKALGLDKK-----VKEPAPTLTTGESKPH 1653
                      *:*********** **::*****.**:*:**  ***        :** :*   :.*.* *

SP|Q6UB99|ANR11_HUMAN PASGADSKDWLAGPHMKEVLPASPRPDQSRPTGVPTPTSVLSCPSYEEVMHTPRTPSCSA 1729
SP|E9Q4F7|ANR11_MOUSE SGPGTESKDWLSGQPLKEVLPASPRTEQSRPTGVPTPTSVVSCPSYEEVMHTPRTPSCSA 1713
                        . *::*****:*  :********* :*************:*******************

SP|Q6UB99|ANR11_HUMAN DDYADLVFDCADSQHSTPVPTAPTSACSPSFFDRFSVASSGLSENA-SQAPARPLSTNLY 1788
SP|E9Q4F7|ANR11_MOUSE DDYPDLVFDCTDSQHSMPVSTASTSACSPPFFDRFSVASSVVSENAAGQTPTRPISTNLY 1773
                      *** ******:***** ** ** ****** ********** :****  .*:*:**:*****

SP|Q6UB99|ANR11_HUMAN RSVSVDIRRTPEEEFSVGDKLFRQQSVPAASSYDSPMPPSMEDRAPLPPVPAEKFACLSP 1848
SP|E9Q4F7|ANR11_MOUSE RSISVDIRRTPEEEFSAGDKLFRQQSVPAPSSFDSPVQHLLEEKAPLPPVPAEKFACLSP 1833
                      **:*************.************ **:***:    :*::****************

SP|Q6UB99|ANR11_HUMAN GYYSPDYGLPSPKVDALHCPPAAVVTVTPSPEGVFSSLQAKPSPSPRAELLVPSLEGALP 1908
SP|E9Q4F7|ANR11_MOUSE GYYSPDYGIPSPKVDTLHCPPTAVVSATPPPDSVFSNLPPKSSPSPRGELLSPAIEGTLP 1893
                      ********:******:*****:***:.** *:.***.*  * *****.*** *::**:**
```

```
SP|Q6UB99|ANR11_HUMAN PDLD----TSEDQQATAAIIPPEPSYLEPLDEGPFSAVITEEPVEWAHPSEQ--ALASSL 1962
SP|E9Q4F7|ANR11_MOUSE PDLGLPLDATEDQQATAAILPQEPSYLEPLDEGPFTTVITEEPVEWTHTAAEQGLSSSL 1953
                      ***.    ::*********:* *************::*********:* : :    :***

SP|Q6UB99|ANR11_HUMAN IGGTSENPVSWPVGSDLLLKSPQRFPESPKRFCPADPLHSAAPGPFSASEAPYPAPPASP 2022
SP|E9Q4F7|ANR11_MOUSE IASASENPVSWPVGSELMLKSPQRFAESPKHFCPGESLHSTTPGPYSAAEPTYPV---SP 2010
                      *..:***********:*:******* ****:***.: ***::***:**:*  **.    **

SP|Q6UB99|ANR11_HUMAN APYALPVAEPGLEDVKDGV-DAVPAAIST-SEAAPYAPPSGLESFFSNCKSLPEAPLDVA 2080
SP|E9Q4F7|ANR11_MOUSE GSYPLPAPEPALEEVKDGGTGAIPVAISAAEGAAPYAAPARLESFFSNCKSHPDAPLDTA 2070
                      . * **. **.**:****  .*:*.***: . ***** *: ********** *:****.*

SP|Q6UB99|ANR11_HUMAN PEPACVAAVAQVEALGPLENSFLDGSRGLSHLGQVEPVPWADAFAGPEDDLDLGPFSLPE 2140
SP|E9Q4F7|ANR11_MOUSE PEPTGVTAVAQVEALGPLESSFLDSNPSISTLSQVEPVSWHEAFTSPEDDLDLGPFSLPE 2130
                      ***: *:***********.****.. .:* *.***** * :**:.**************

SP|Q6UB99|ANR11_HUMAN LPLQTKDAADGEAEPVEESLAPPEEMPPGAPGVINGGDVSTVVAEEPPALPPDQASTRLP 2200
SP|E9Q4F7|ANR11_MOUSE LPLQAKDASDVEAEAAKASPVPPAESPPGPTGVLGGGDVPAPAAEEPPAPPPQEASPQLS 2190
                      ****:***:* *** .: * .** * ***  **:.**** : .****** **::** :*

SP|Q6UB99|ANR11_HUMAN AELEPEPSGEPKLDVALEAAVEAETVPEERARGDPDSSVEPAPVPPEQRPLGSGDQGAEA 2260
SP|E9Q4F7|ANR11_MOUSE --TEPEPSEEPKLDVVLEATVETEVLADDSAPEASISNSVPAPSPPQQQPPGGGDEEAET 2248
                        ***** ******.***:**:*.: :: *     *.  *** **:*:* *.**: **:

SP|Q6UB99|ANR11_HUMAN EGPPAASLCAPDGPAPNTVAQAQAADGAGPEDDTEASRAAAPAEGPPGGIQPEAA--EPK 2318
SP|E9Q4F7|ANR11_MOUSE EDPSATPCCAPDGPTTDGLAQAHN--------SAEASCVVAAAEGPPGNVQAEATDPEPK 2300
                      *.* *:  ******: : :***:         .:*** ..* ******.:* **:  ***

SP|Q6UB99|ANR11_HUMAN PTAEAPKAPRVEEIPQRMTRNRAQMLANQSKQGPPPSEKECAPTPAPVTRAKARGSEDDD 2378
SP|E9Q4F7|ANR11_MOUSE PTSEVPKAPKVEEVPQRMTRNRAQMLASQSKQGIPAAEKDP--MPTPASRAKGRASEEED 2358
                      **:*.****:***:*************.***** * :**:    *:*.:***.*.**::*

SP|Q6UB99|ANR11_HUMAN AQAQHPRKRRFQRSTQQLQQQLNTSTQQTREVIQQTLAAIVDAIKLDAIEPYHSDRANPY 2438
SP|E9Q4F7|ANR11_MOUSE AQAQHPRKRRFQRSSQQLQQQLNTSTQQTREVIQQTLAAIVDAIKLDAIEPYHSDRSNPY 2418
                      **************:*****************************************:***

SP|Q6UB99|ANR11_HUMAN FEYLQIRKKIEEKRKILCCITPQAPQCYAEYVTYTGSYLLDGKPLSKLHIPVIAPPPSLA 2498
SP|E9Q4F7|ANR11_MOUSE FEYLQIRKKIEEKRKILCCITPQAPQCYAEYVTYTGSYLLDGKPLSKLHIPVIAPPPSLA 2478
                      ************************************************************

SP|Q6UB99|ANR11_HUMAN EPLKELFRQQEAVRGKLRLQHSIEREKLIVSCEQEILRVHCRAARTIANQAVPFSACTML 2558
SP|E9Q4F7|ANR11_MOUSE EPLKELFKQQEAVRGKLRLQHSIEREKLIVSCEQEILRVHCRAARTIANQAVPFSACTML 2538
                      *******:****************************************************

SP|Q6UB99|ANR11_HUMAN LDSEVYNMPLESQGDENKSVRDRFNARQFISWLQDVDDKYDRMKTCLLMRQQHEAAALNA 2618
SP|E9Q4F7|ANR11_MOUSE LDSEVYNMPLESQGDENKSVRDRFNARQFISWLQDVDDKYDRMKTCLLMRQQHEAAALNA 2598
                      ************************************************************

SP|Q6UB99|ANR11_HUMAN VQRMEWQLKVQELDPAGHKSLCVNEVPSFYVPMVDVNDDFVLLPA 2663
SP|E9Q4F7|ANR11_MOUSE VQRMEWQLKAQELDPAGHKSLCVNEVPSFYVPMVDVNDDFVLLPA 2643
                      *********.***********************************
```

#### Figure S3: Four variants are located at three residues in predicted destruction motifs

ANKRD11 amino acid sequence with the variants identified in affected individuals shaded in yellow. Different degradation motifs are indicated: RxxL-motifs are underlined, Proviz-predicted destruction motifs are bold, with D-boxes in red, Ken-boxes in green and an Abba-motif in orange.

>sp|Q6UB99|ANR11_HUMAN Ankyrin repeat domain-containing protein 11 OS=Homo sapiens OX=9606 GN=ANKRD11 PE=1 SV=3

MPKGGCPKAPQQEELPLSSDMVEKQTGKKDKDKVSLTKTPKLERGDGGKEVRERASKRKL
PFTAGANGEQKDSDTEKQGPERKRIKKEPVTRKAGLLFGMGLSGIRAGYPLSERQQVALL
MQMTAEESANSPVDTTPKHPSQSTVCQKGTPNSASKTKDKVNKRNERGETRLHRAAIRGD
ARRIKELISEGADVNVKDFAGWTALHEACNRGYYDVAKQLLAAGAEVNTKGLDDDTPLHD
AANNGHYKVVKLLLRYGGNPQQSNRKGETPLKVANSPTMVNLLLGKGTYTSSEESSTESS
EEEDAPSFAPSSSVDGNNTDSEFEKGLKHKAKNPEPQKATAPVKDEYEFDEDDEQDRVPP
VDDKHLLKKDYRKETKSNSFISIPKMEVKSYTKNNTIAPKKASHRILSDTSDEEDASVTV
GTGEKLRLSAHTILPGSKTREPSNAKQQKEKNKVKKKRKKETKGREVRFGKRSDKFCSSE
SESESSESGEDD**RDSL**GSSGCLKGSPLVLKDPSLFSSLSASSTSSHGSSAAQKQNPSHTD
QHTKHWRTDNWKTISSPAWSEVSSLSD**STRTRLTSESDY**SSEGSSVESLKPVRKRQEH**RK**
**RASLSEKKSP**FLSSAEGAVPKLDKEGKVVKKHKTKHKHKNKEKGQCSISQELKLKSFTYE
YEDSKQKSDKAILLENDLSTENKLKVLKHDRDHFKKEEKLSKMKLEEKEWLFKDEKSLKR
IKDTNKDISRSFREEKDRSNKAEKERSLKEKSPKEEKLRLYKEERKKKSKDRPSKL**EKKN**
**DLK**EDKISKEKEKIFKEDKEKLKKEKVYREDSAFDEYCNKNQFLENEDT**KFSLSDD**QRDR
WFSDLSDSSFDFKGEDSWDSPVTDYRDMKSDSVAKLILETVKEDSKERRRDSRAREKRDY
REPFFRKKD**RDYL**DKNSEKRKEQTEKHKSVPGYLSEKDKKRRESAEAGRDRKDALESCKE
RRDGRAKPEEAH**REEL**KECGCESGFKDKSDGDFGKGLEPWERHHPAREKEKKDGPDKERK
EKTKPERYKEKSSDKDKSEKSILEKCQKDKEFDKCFKEKKDTKEKHKDTHGKDKERKASL
DQGKEKKEKAFPGIISEDFSEKKDDKKGKEKSWYIADIFTDESEDDRDSCMGSGFKMGEA
SDLPRTDGLQEKEEGREAYASDRHRKSSDKQHPERQKDKEPRDRRKDRGAADAGRDKKEK
VFEKHKEKKDKESTEKYKDRKDRASVDSTQ**DKKNKQK**LPEKAEKKHAAEDKAKSKHKEKS
DKEHSKERKSSRSADAEKSLLEKLEEEALHEYREDSNDKISEVSSDSFTDRGQEPGLTAF
LEVSFTEPPGDDKPRESACLPEKLKEKERHRHSSSSSKKSHDRERAKKEKAEKKEKGEDY
KEGGSRKDSGQYEKDFLEADAYGVSYNMKADIEDELDKTIELFSTEKKDKNDSEREPSKK
IEKELKPYGSSAINILKEKKKREKHREKWRDEKERHRDRHADGLLRHH**RDEL**LRHHRDEQ
KPATRDKDSPPRVLKDKSRDEGPRLGDAKLKEKFKDGAEKEKGDPVKMSNGNDKVAPSKD
PGKKDARP**REKL**LGDGDLMMTSFERMLSQKDLEIEERHKRHKERMKQMEKLRHRSGDPKL
KEKAKPADDGRKKGLDIPAKKPPGLDPPFKDKKLKESTPIPPAAENKLHPASGADSKDWL
AGPHMKEVLPASPRPDQSRPTGVPTPTSVLSCPSYEEVMHTPRTPSCSADDYADLVFDCA
DSQHSTPVPTAPTSACSPSFFDRFSVASSGLSENASQAPARPLSTNLYRSVSVDIRRTPE
EEFSVGDKLFRQQSVPAASSYDSPMPPSM**EDRAPLPPVPAE**KFACLSPGYYSPDYGLPSP
KVDALHCPPAAVVTVTPSPEGVFSSLQAKPSP**SPRAELLVPSLE**GALPPDLDTSEDQQAT
AAIIPPEPSYLEPLDEGPFSAVITEEPVEWAHPSEQALASSLIGGTSENPVSWPVGSDLL
LKSPQRFPESPKRFCPADPLHSAAPGPFSASEAPYPAPPASPAPYALPVAEPGLEDVKDG
VDAVPAAISTSEAAPYAPPSGLESFFSNCKSLPEAPLDVAPEPACVAAVAQVEALGPLEN
SFLDGSRGLSHLGQVEPVPWADAFAGPEDDLDLGPFSLPELPLQTKDAADGEAEPVEESL
APPEEMPPGAPGVINGGDVSTVVAEEPPALPPDQASTRLPAELEPEPSGEPKLDVALEAA
VEAETVPEERARGDPDSSVEPAPVPPEQRPLGSGDQGAEAEGPPAASLCAPDGPAPNTVA
QAQAADGAGPEDDTEASRAAAPAEGPPGGIQPEAAEPKPTAEAPKAPRVEEIPQRMTRNR
AQMLANQSKQGPPPSEKECAPTPAPVTRAKARGSEDDDAQAQHPRKRRFQRSTQQLQQQL
NTSTQQTREVIQQTLAAIVDAIKLDAIEPYHSDRANPYFEYLQIRKKIE**EKRKILCCITP**
**Q**APQCYAEYVTYTGSYLLDGKPLSKLHIPVIAPPPSLAEPLKELFRQQEAV**RGKL**RLQHS
**IEREKLIVSCEQ**EILRVHCRAARTIANQAVPFSACTMLLDSEVYNMPLESQ**GDENKSVR**D
RFNARQFISWLQDVDDKYDRMKTCLLMRQQHEAAALNAVQRMEWQLKVQELDPAGHKSLC
VNEVPSFYVPMVDVNDDFVLLPA

#### Figure S4: *ANKRD11* missense variants affecting arginine residues in RD2 are overrepresented in the cohort

The observed number of mutated arginine residues in our cohort (12/17 RD2; 0/8 outside RD2) were compared against an expected distribution of mutated arginine residues in ANKRD11 (see Methods). The mean (circle) and standard deviation (interval) of the expected distribution is shown in red. The black diamond represents the observed distribution inside RD2 (top) and outside (bottom). Permutation *p*-values shown above the expected distribution represent the likelihood that the observed distribution would occur by chance.

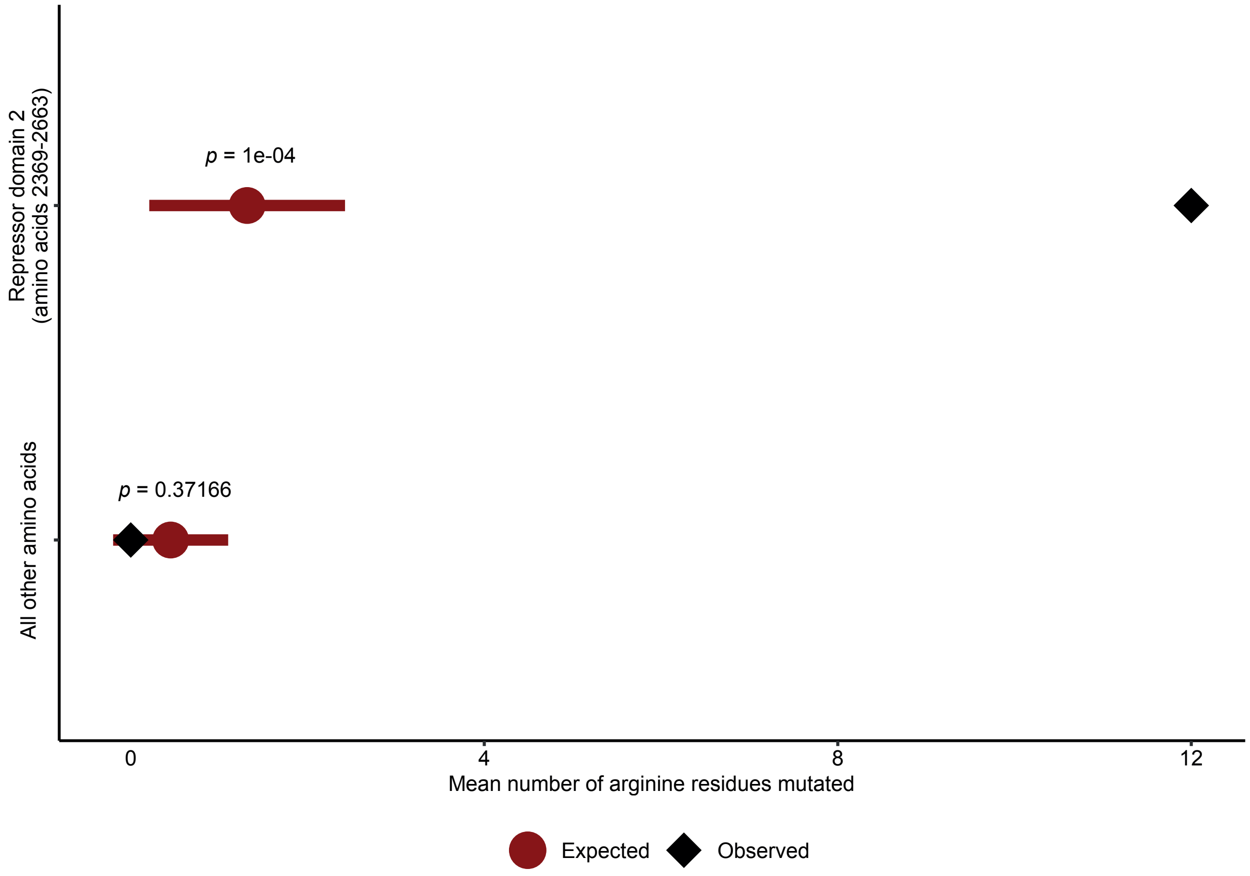

#### Figure S5: Quantification of ANKRD11 nuclear speckles (related to Figure 3)

Results of quantification of (A) number, (B) size, (C) perimeter, (D) area covered, (E) solidity and (F) and density of ANKRD11 nuclear speckles. Values are expressed relative to wildtype (WT) and represent the mean ± SEM of three independent experiments (one-way ANOVA and a post-hoc Dunnett's test).

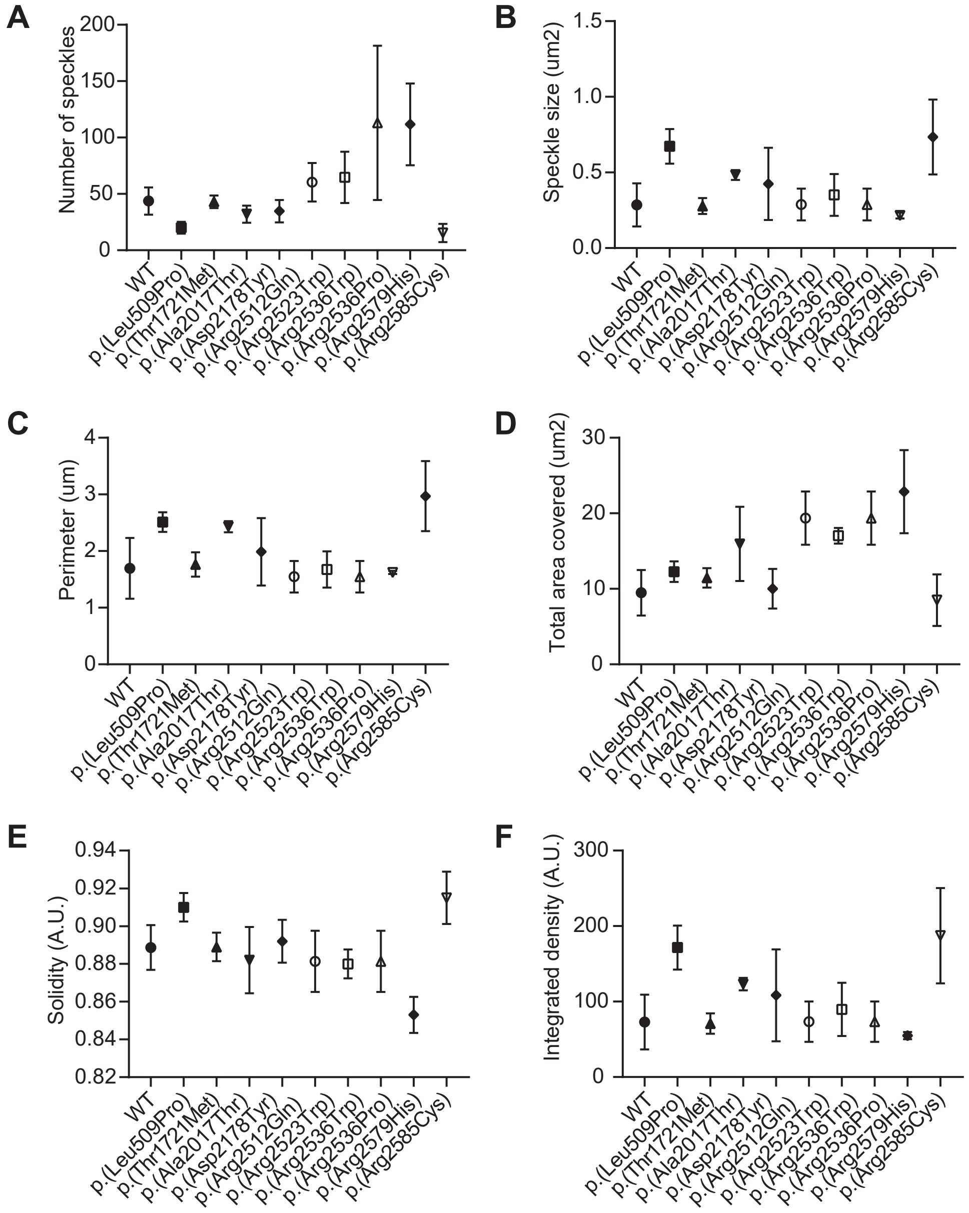

#### Figure S6: EGFP-ANKRD11 protein expression in transiently transfected HEK293T/17 cells

Immunoblot of whole cell lysates of HEK293T/17 cells expressing EGFP-tagged ANKRD11 variants probed with anti-GFP antibodies. β-actin was used as a loading control.

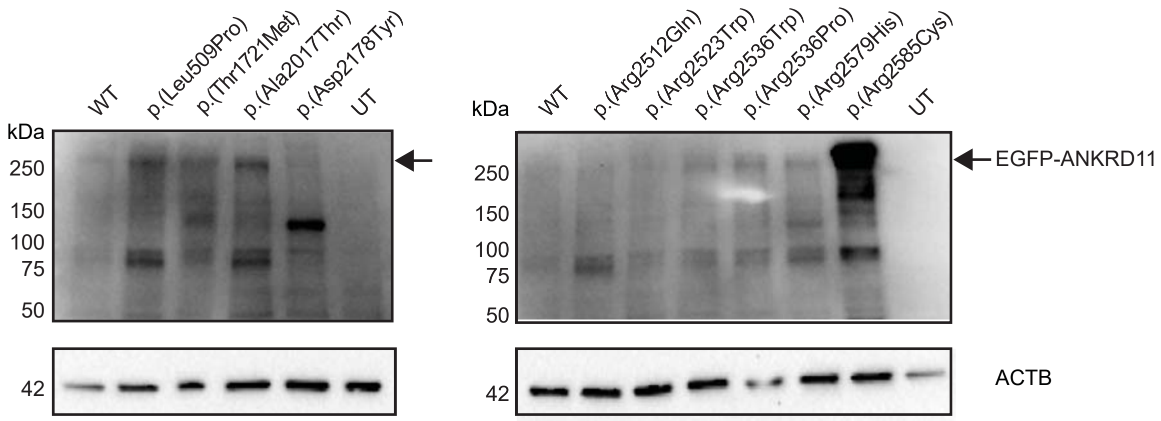

### Figure S7: Stability and degradation of ANKRD11 variants as EGFP-fusion protein in HEK293T/17 cells

Relative expression ANKRD11 variants as EGFP-fusion protein in HEK293T/17 cells treated with (A) 50µg/ml cycloheximide (CHX) or (B) 5µg/ml proteasome inhibitor MG132. Equal volume of DMSO was used as a vehicle control. Fluorescence intensity was measured for 24 hours with three-hour intervals. Values are expressed relative to t= 0 hour and represent the mean ± SEM of three independent experiments, each preformed in triplicates (*p<0.05, **p<0.01, ***p<0.001, ****p<0.0001 CHX or MG132 versus DMSO; repeated measure two-way ANOVA and a post-hoc Sidak's test).

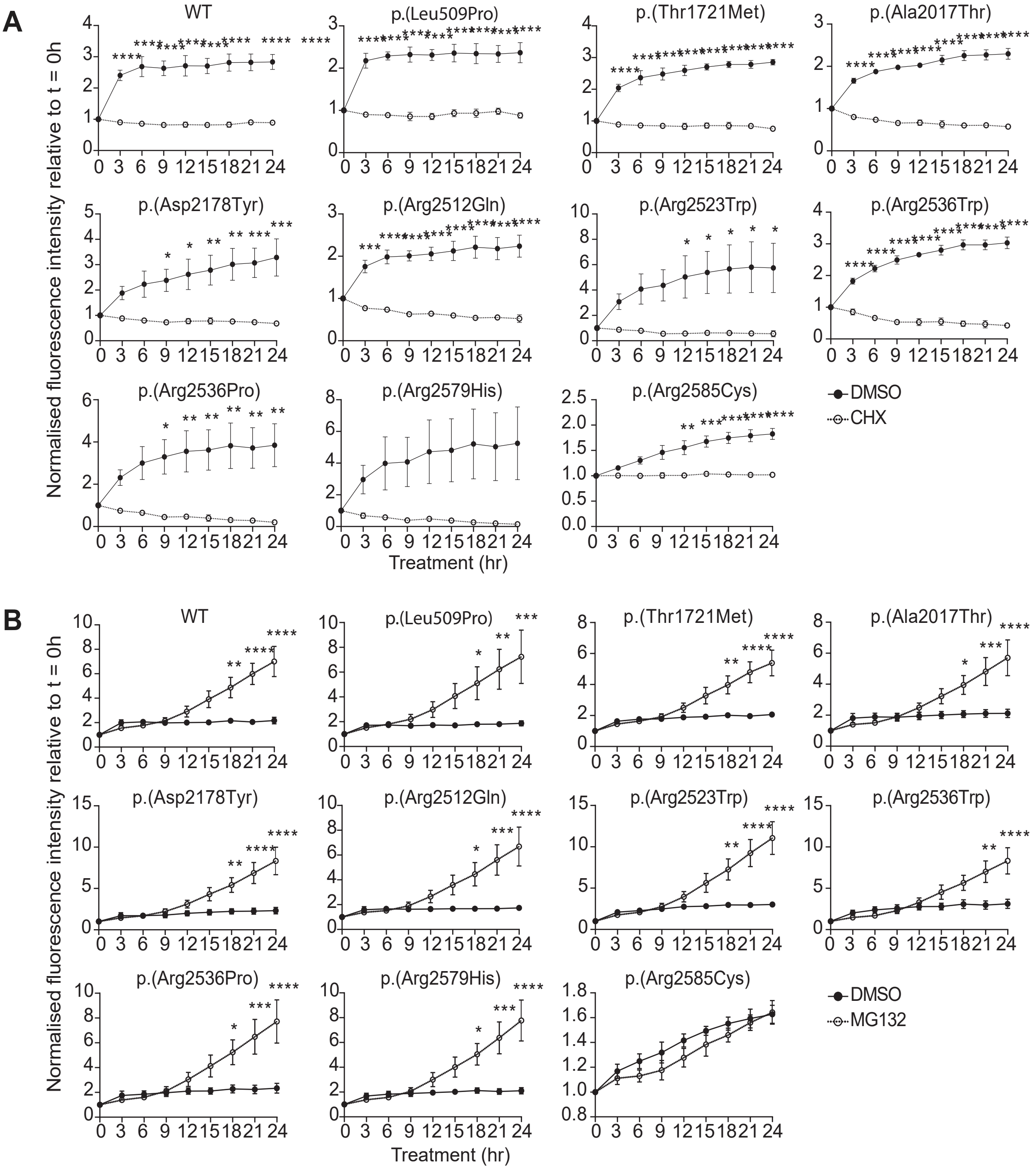

### Figure S8: Potential effects on cryptic splice sites

Alamut splice site tools predict potential effects of p.(Gly1093Arg) and p.(Asp2178Tyr), respectively predicted to remove a cryptic acceptor splice site, and to introduce a cryptic donor splice site.

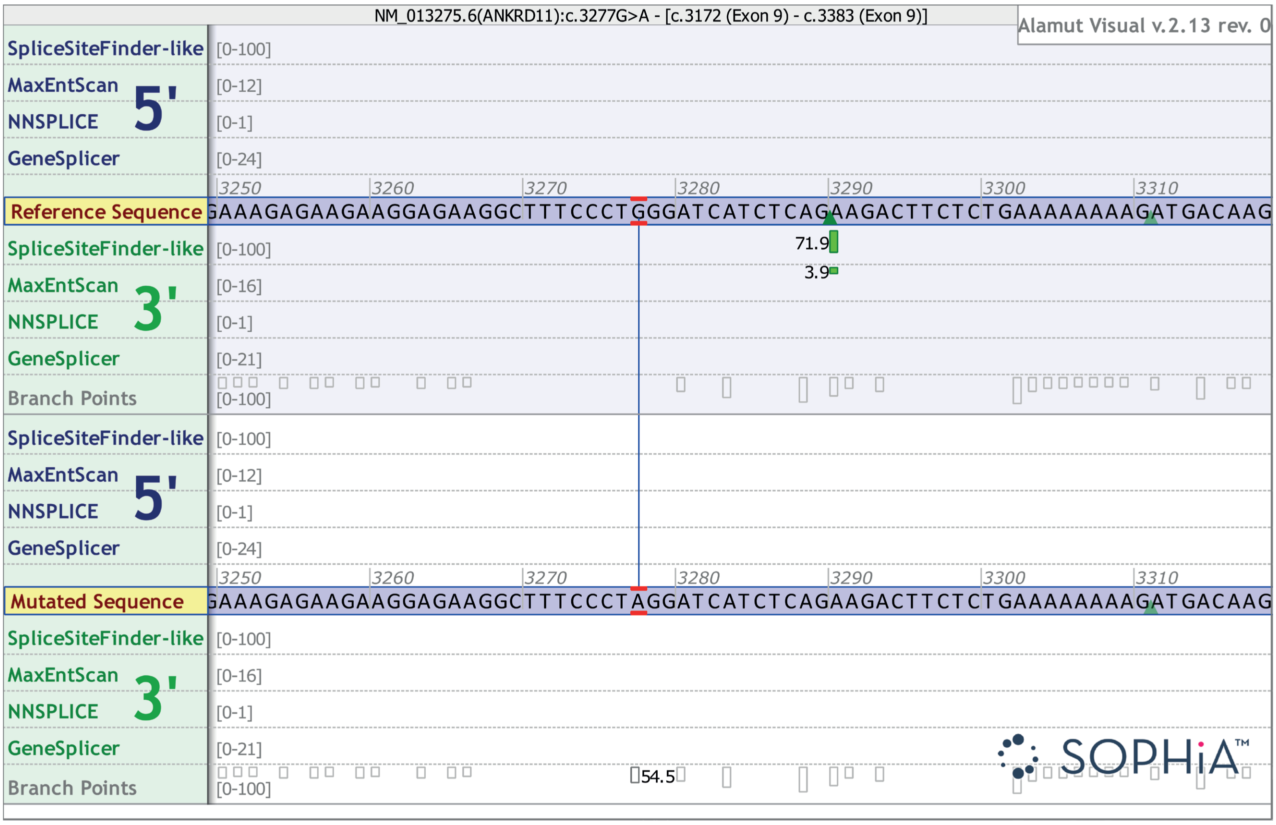


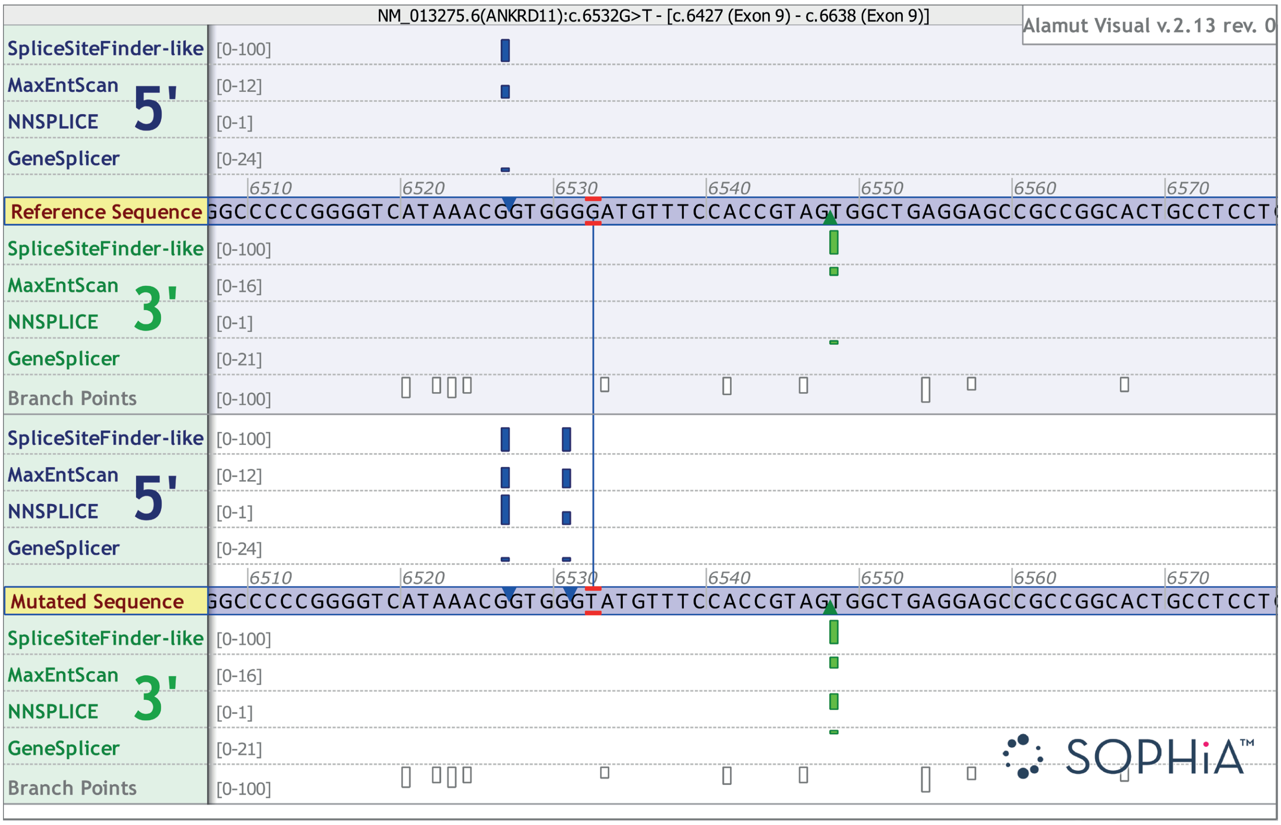
