## Supplementary information for "Missense variants in *ANKRD11* cause KBG syndrome by impairment of stability or transcriptional activity of the encoded protein"

**Page 1: Overview of all supplementary files**

**Page 2: Supplementary information related to the introduction**

- ANKRD11 missense variants reported in literature

**Page 2: Supplementary methods**

- Identification of *ANKRD11* variants, clinical characterization and in silico predictions
- Human Phenotype Ontology (HPO)-based phenotype clustering analysis
- Data of individuals obtained from the Radboudumc Biobank and Radboudumc KBG

national referral centre

- Spatial clustering analysis of missense variants
- Immunoblotting

**Page 5: Supplementary results**

- ANKRD11 missense variants cause syndromic neurodevelopmental phenotypes

**Page 6: Supplementary references**

**Supplementary files (provided separately):**

- Table S1: ANKRD11 missense variants reported in literature
- Table S2: Clinical information per individual – not included in this preprint, available from the corresponding author on request
- Table S3: Simplified clinical features based on HPO-data using the Wang algorithm and clustering of individuals
- Table S4: List of primers & antibodies
- Table S5: List of variants, including annotations and classification based on ACMG
- Table S6: Destruction motifs
- Figure S1: Pedigree of the family with five affected individuals carrying p.(Arg2579His) – not included in this preprint, available from the corresponding author on request
- Figure S2: Variant p.(Glu2522Lys) in individuals 13 and 14 is equivalent to Yoda variant p.(Glu2502Lys)
- Figure S3: Four variants are located at 3 residues in predicted destruction motifs
- Figure S4: *ANKRD11* missense variants affecting arginine residues in RD2 are overrepresented in the cohort
- Figure S5: Quantification of ANKRD11 nuclear speckles.
- Figure S6: EGFP-ANKRD11 protein expression in transiently transfected HEK293T/17 cells.
- Figure S7: Relative expression ANKRD11 variants as EGFP-fusion protein in HEK293T/17 cells treated with (A) 50µg/ml cycloheximide (CHX) or (B) 5µg/ml proteasome inhibitor MG132.
- Figure S8: Potential effects on cryptic splice sites
- Supplementary_HPO-data_in_JSON – not included in this preprint, available from the corresponding author on request

**SUPPLEMENTARY INFORMATION RELATED TO INTRODUCTION**

***ANKRD11* missense variants reported in literature**

Individuals with *ANKRD11* missense variants are listed in Table S1 [1-20].

**SUPPLEMENTARY METHODS**

**Identification of *ANKRD11* variants, clinical characterization and *in silico* predictions**

Individuals with (likely) pathogenic *ANKRD11* missense variants were identified through international collaborations facilitated by MatchMaker Exchange [21], the Decipher Database [22], the Solve-RD consortium and RD-connect [23]. Variants were identified by WES or Sanger sequencing as previously described [24, 25]. We annotated variants to NM_013275.6 in GRCh37/Hg19. Pathogenicity of variants was assessed by the following *in silico* tools: CADD-PHRED V1.6 [26], PhyloP 2015 [27], GERP 2013 [28], Align GVGD v2007 [29], SIFT v6.2.0 [30], MutationTaster v2013 [31], Grantham [32], Metadome [33] and SpliceAI [34]. The presence of degron motifs was analysed with ProViz [35]. Variants were classified according to ACMG guidelines [36]. Individuals were clinically characterized by reviewing medical files, and/or revising the phenotypes in outpatient clinics. Phenotypic fit to the KBG-associated clinical spectrum was assessed by two medical geneticists with expertise in KBG syndrome, based on diagnostic criteria [37] and frequently described features in four large cohorts [37-40]. We obtained informed consent to publish unidentifiable data for all individuals reported in this study. Specific consent was obtained for publication of clinical photographs. Consent procedures were in accordance with the Declaration of Helsinki and local ethical guidelines of participating centres. Consent procedures and details of the IRB/oversight body that provided approval or exemption for the research described are described in the ethical statement. Figure 2, Table S2, Figure S1 and Supplementary JSON are not included in this preprint and are available from the corresponding author on request.

**Human Phenotype Ontology (HPO)-based phenotype clustering analysis**

To quantify potential differences between phenotypic features of individuals with ANKRD11 missense variants and individuals with ANKRD11 PTVs or microdeletions, HPO-based clustering analysis was performed as previously described [41]. Clinical data of 29 individuals carrying ANKRD11 missense variants (Table S2, Supplementary JSON), and of 30 individuals with KBG syndrome caused by PTVs or 16q24 microdeletions obtained from the Radboudumc expert centre for rare neurodevelopmental disorders via Biobank Genetics and Rare Disease were standardized using HPO terminology (release 2018-12-21) [42] in PhenoTips (<https://github.com/phenotips/phenotips>, version 1.4.1) [43]. To avoid interobserver bias, all standardization of clinical data to HPO terminology was performed by the same clinician. In brief, semantic similarity between all HPO terms was calculated with the Wang algorithm in the HPOSim R-package [44, 45]. Terms were grouped and replaced by an overarching new feature when having a similarity score ≥0.5 (Table S3). To quantify a potential difference between cases with missense variants and cases with PTVs/microdeletions, we used Partitioning Around Medoids (PAM) clustering [46] dividing the cohort into two groups, followed by a permutation test (100,000x). p-values <0.05 were considered significant.

**Data of individuals obtained from the Radboudumc Biobank and Radboudumc KBG national referral centre**

To compare the individuals with ANRKD11 missense variants observed in the cohort to KBG syndrome resulting from PTVs and 16q24.3 microdeletions, we used information on genotype and phenotype of 30 individuals diagnosed with KBG syndrome obtained from the Radboudumc Biobank Genetics and Rare Disease (<https://www.radboudumc.nl/en/research/radboud-technology-centers/radboud-biobank>) and the Radboudumc KBG national referral centre. The complete HPO-data and genotypic information of these individuals are not shared as a supplementary file accompanying this manuscript, due to IRB and General Data Protection Regulation (EU GDPR) restrictions. Access to this data may be requested from the data availability committee by contacting the corresponding author.

The 30 individuals obtained from the Radboudumc Biobank are a representative subset of all individuals with KBG syndrome that are in clinical care of the Radboudumc KBG national referral centre (<https://www.radboudumc.nl/en/centers-of-clinical-expertise/centers-of-clinical-expertise-for-rare-diseases/rare-congenital-developmental-disorders>), capturing the full phenotypic spectrum of KBG syndrome, including mild and subclinical presentations. Of the 30 individuals, 17 were male and 13 female, with an age range of 3 to 73 years. Genotypically, three individuals had a 16q24.3 deletion, 20 individuals carried frameshift variants, and seven individuals had a nonsense variant. Of the 27 single nucleotide variants and indels, 26 located to the long exon 9, introducing a premature stop codon that is predicted to result in escape from the nonsense-mediated decay (NMD) pathway, whereas only one variant is predicted to trigger NMD [47, 48]. Variants were shown to be *de novo* in 18 individuals. For four individuals, the variant occurred in a familial context, and for the final eight, inheritance could not be established.

**Spatial clustering analysis of missense variants**

25 of the 29 observed missense variants were included in spatial clustering analysis, after removal of four variants because of familial occurrence. The geometric mean was computed over the locations of observed missense variants in the cDNA of ANKRD11 (7,992 bp) and subsequently compared to each of the geometric means of 1,000,000 permutations of randomly redistributing the variant locations over the total coding sequence of ANKRD11. A *p*-value was obtained by calculating how often the observed geometric distance was smaller than the permutated geometric mean distance [49, 50] and considered significant if <0.05.

**Immunoblotting**

Whole-cell lysates were prepared as described previously [51]. Total protein was quantified using the Pierce BCA protein assay kit (Thermo Fisher). Proteins were resolved on 4–15% Tris-Glycine gels and transferred to PDVF membranes (Bio-Rad). After blotting, membranes were incubated overnight at 4˚C with the appropriate primary antibodies. Membrane were then incubated with HRP-conjugated secondary antibodies. Proteins were visualized using the Novex ECL Chemiluminescent Substrate Reagent kit (Invitrogen) or SuperSignal West Femto Maximum Sensitivity Substrate (Thermo Fisher) and the ChemiDoc XRS+ System (Bio-Rad). A list of antibodies used can be found in Table S4.

**SUPPLEMENTARY RESULTS**

***ANKRD11* missense variants cause syndromic neurodevelopmental phenotypes**

In addition to the most frequently observed phenotypic features described in the manuscript, individuals presented with a variety of additional features (summarized in Table 1 with details provided in Table S2). We observed seizures in six cases (6/27, 22.2%), ranging in severity from absence seizures to refractory complex focal seizures. Additional neurological features were predominantly mild, with sleep disturbances (8/26, 30.8%) and hypotonia (10/24, 41.7%) most frequently reported. Commonly observed behavioral problems included ADHD or hyperactivity (18/26, 69.2%), ASD (9/25, 36%) and anxiety (9/24, 37.5%), together with a spectrum of additional behavioral symptoms. Although most individuals were born at term (23/24, 95.8%), both pre- and perinatal complications were prevalent (11/24, 45.8% and 15/26, 57.7% respectively), including maternal pregnancy-related illness, a variety of ultrasound abnormalities, complicated delivery and asphyxia. Short stature was seen in over half of the cohort (15/28, 53.6%), explained by growth hormone deficiency in only two individuals, and seven cases presented with abnormal head circumference (macrocephaly 1/27, 3.7%; microcephaly 6/27, 22.2%). Congenital heart defects occurred in 32% of individuals (8/25), mostly consisting of atrial and/or ventricular septal defects and cardiac valve abnormalities. Cryptorchidism was observed in 20% of male individuals (3/15), and other urogenital abnormalities included duplication of the kidney or renal collecting system and hypospadias. The most prominent problems of the gastrointestinal tract were feeding difficulties (9/27, 33.3%), constipation (6/27, 22.2%) and gastroesophageal reflux disease (3/27, 11.1%). Over half of all individuals showed delayed bone maturation (8/14, 57.1%), but other abnormalities of the skeletal system were not frequently reported. Abnormalities of vision occurred in 50% (13/26) of individuals, comprising hypermetropia, myopia, astigmatism and strabismus. Hearing loss, resulting from recurrent or chronic otitis media, stapes ankylosis or ear atelectasis, was also prevalent (11/28, 39.3%). Only two individuals had a cleft palate, and two individuals were affected by velopharyngeal insufficiency. Additionally, a wide range of other abnormalities was observed at low frequencies (Table S2), most remarkably choanal atresia (individual 4) and ileal atresia (individual 19).
